# Non-occupational physical activity and risk of 22 cardiovascular disease, cancer, and mortality outcomes: a dose-response meta-analysis of large prospective studies

**DOI:** 10.1101/2022.03.02.22271753

**Authors:** Leandro Garcia, Matthew Pearce, Ali Abbas, Alexander Mok, Tessa Strain, Sara Ali, Alessio Crippa, Paddy C Dempsey, Rajna Golubic, Paul Kelly, Yvonne Laird, Eoin McNamara, Samuel Moore, Thiago Herick de Sa, Andrea D Smith, Katrien Wijndaele, James Woodcock, Søren Brage

## Abstract

**Objective:** To estimate dose-response associations between non-occupational physical activity and multiple chronic disease outcomes in the general adult population.

**Eligibility criteria:** Prospective cohort studies with (a) general population samples >10,000 adults, (b) ≥3 exposure categories, and (c) risk measures and confidence intervals for all-cause mortality, total cardiovascular disease, coronary heart disease, stroke, heart failure, total cancer, and site-specific cancers (head and neck, myeloid leukemia, myeloma, gastric cardia, lung, liver, endometrium, colon, breast, bladder, rectum, esophagus, prostate, kidney).

**Information sources:** PubMed, Scopus, Web of Science, and reference lists of published studies, searched in February 2019.

**Data extraction and synthesis:** Independent extraction and double-checking of study characteristics, exposure, and outcome assessment by two reviewers for each paper. Primary exposure was non-occupational physical activity volume, harmonized to physical activity energy expenditure in marginal MET-hours per week (mMET-h/week). The current minimum recommendations for physical activity (150 min/week of moderate-to-vigorous physical activity) equate to 8.75 mMET-h/week. Outcomes were risks of mortality, cardiovascular diseases, and cancers. We used restricted cubic splines in random-effects meta-analyses. Potential population impact was quantified using impact fractions.

**Results:** 196 articles were included, covering 94 cohorts. The evidence base was largest for all-cause mortality (50 independent results; 163,415,543 person-years; 811,616 events), and incidence of cardiovascular disease (37 independent results; 28,884,209 person-years; 74,757 events) and cancer (31 independent results; 35,500,867 person-years; 185,870 events). In general, inverse non-linear associations were observed, steeper between 0 and 8.75 mMET-h/week, with smaller marginal reductions in risk above this level to 17.5 mMET-h/week, beyond which additional reductions were small and uncertain. Associations were stronger for all-cause and cardiovascular disease mortality than for cancer mortality. If all insufficiently active individuals had met the recommended physical activity level, 15.7% (95%CI: 13.1 to 18.2%) of all premature deaths would have been averted.

**Conclusions:** Inverse non-linear dose-response associations suggest substantial protection against a range of chronic disease outcomes from small increases in non-occupational physical activity in inactive adults.

**Review registration:** PROSPERO CRD42018095481.

## 1. INTRODUCTION

Cardiovascular disease (CVD) is the leading cause of death globally, responsible for 17.9 million annual deaths in 2019,^1^ whereas cancers top the global disease burden with respect to disability-adjusted life-years.^2^ The relative contribution of each risk and protective factor to the incidence of, and mortality from, these conditions is an ongoing debate.

Higher levels of physical activity (PA) have been associated with lower rates of premature mortality and chronic disease outcomes.^3^ However, the shape of the dose-response association has been more difficult to determine, and is yet to be established for a range of diseases. One reason is that more data are needed to examine a wide enough exposure range and less common outcomes; another is that large population studies use a variety of instruments to assess PA. While this makes the task of conducting a dose-response meta-analysis harder, it is possible through exposure harmonization.^4, 5^

Accurate estimation of the dose-response association between PA and disease outcomes, combined with prevalence estimates, is a prerequisite for assessing the population disease burden of insufficient PA and the potential impact of changes to population levels of PA. Modelling studies that estimate the impact of these strategies have found that the shape of the dose-response association makes a significant difference when quantifying population health impacts.^6^

Pooled analyses of the association between leisure-time PA and mortality have been undertaken in six US studies, showing an inverse, curvilinear dose-response association,^7^ but the totality of evidence has not recently been assessed. For cancer and CVD outcomes, the Global Burden of Disease (GBD) Study reported non-linear dose-response associations for total PA (i.e., including physical activity performed during work, or occupational activity), estimating that a dose above 65 MET-h/week is associated with a 20% risk reduction.^5^ The occupational component of PA is often crudely measured via self-report or not measured at all and there is some evidence that it might have a different relation to health endpoints compared with leisure-time PA.^8-10^

Using extensive exposure harmonization to quantify non-occupational PA, we therefore examined the dose-response association between non-occupational PA and the risk of 22 CVD, cancer, and mortality outcomes. To contextualize our results for public health promotion, we used potential population impact fractions to estimate the proportion of preventable deaths and disease outcomes at different non-occupational PA levels.

## 2. METHODS

The study protocol is available at PROSPERO, registration number CRD42018095481.

### 2.1 Eligibility criteria

Supplementary eMethods 1 shows the study inclusion and exclusion criteria. We included prospective cohort studies that followed adults (≥ 18 years) without pre-existing conditions, reported PA at baseline in at least three ordinal exposure levels, and reported risk estimates for the examined outcomes. We excluded studies with < 10,000 participants (to limit potential bias from small study sizes with positive results) or if the follow-up period was < 3 years (to minimize reverse causality bias).

Only articles that examined leisure-time PA, alone or in combination with other domains or specific types of activity, were included. We excluded articles whose measures of PA contained occupational activity that could not be factored out, and that investigated individual domains of PA that did not include leisure-time activity (e.g., transport-related physical activity only).

The outcomes of interest were:

- All-cause, total CVD, and total cancer mortality;
- Total CVD, coronary heart disease, stroke, and heart failure incidence (fatal and non-fatal);
- Total and site-specific (head and neck, myeloid leukemia, myeloma, gastric cardia, lung, liver, endometrial, colon, breast, bladder, rectum, esophageal, prostate, and kidney) cancer incidence (fatal and non-fatal). These site-specific cancers were selected based on previously reported associations with PA.^11^

### 2.2 Search and selection process

We searched PubMed, Scopus, Web of Science, and reference lists of reviews retrieved from our systematic review, known to the authors, or cited in the 2018 U.S. Physical Activity Guidelines Advisory Committee Scientific Report.^12^ The Supplement (eMethods 2) details the systematic search strategy. We considered peer-reviewed articles in any language published in academic journals until February 2019.

Titles and abstracts and, subsequently, full texts were screened independently twice for eligibility (Supplement, eFigure1). Disagreements were resolved by discussion.

If multiple articles reported results on the same cohort and outcome, we followed specific criteria (eMethods 3) to select one.

### 2.3 Data extraction

Data from each paper were extracted by one reviewer and double-checked independently by a second reviewer. Disagreements were checked against the full text and resolved by discussion.

We extracted data on publication (first author, year of publication), study characteristics (country, cohort name, sample size, age and sex of participants, and duration of follow-up), PA exposure assessment (instrument, domains of activity, and exposure categories), and outcome assessment. We also identified how original study analyses had considered baseline morbidity (exclusion at baseline or statistical adjustment in multivariable regression models) and early incident cases during follow-up.

For each exposure category, we extracted information to quantify PA volume, number of cases, number of participants and/or person-years of follow-up, and risk estimates with 95% confidence intervals (95%CIs). Risk estimates from the most adjusted model were used. When available, risk estimates of the most adjusted model without adiposity-related covariates (e.g., BMI, waist circumference) were retrieved and used in sensitivity analyses. Results reported separately by sex or other attributes (e.g., age, ethnicity) or for multiple cohorts within an article were treated as separate associations. When necessary, data were obtained from other publications using the same cohort, imputation procedures (Supplement, eMethods 4), or by contacting authors.

### 2.4 Physical activity exposure harmonization

#### 2.4.1 Overview of harmonization

We harmonized reported PA exposure levels from all included studies into a common metric of non-occupational PA volume in marginal MET-hours per week (mMET-h/week), reflecting the rate of energy expenditure outside of work, above the resting metabolic rate (1 MET). This allows correct equating of PA volume of time spent at different PA intensity levels.^4^ The principles of each aspect of our comprehensive harmonization procedure are described below and presented in a flowchart available in our <u>OSF repository</u>. The last columns of the “Details of included articles and original and harmonized exposures” table in the same <u>OSF repository</u> contain the details of how the original PA exposure categories of each study were harmonized to mMET-h/week.

#### 2.4.2 Frequency, duration, and intensity assumptions

Studies that described PA exposure as frequency and/or duration were converted to weekly duration. If session duration was not provided, we assumed a duration of 0.75 hour/session (0.5 hour/session in sensitivity analyses). Studies reporting categorical frequency data (e.g., never, sometimes, and often) were converted to weekly duration using assumptions for both frequency (e.g., 0, 2, 5 sessions per week) and duration of sessions. When PA intensity was not explicitly described, we considered the reported activities as light, moderate, or vigorous based on the description and using the Compendium of Physical Activities.^13^ We assigned mMET values of 1.5 for light, 3.5 for moderate and moderate-to-vigorous, and 7.0 for vigorous PA (1 mMET less in sensitivity analyses).

#### 2.4.3 Converting absolute energy expenditure to MET values

For studies reporting energy expenditure without adjustment for body weight (e.g., kJ/day, kcal/week), energy expenditure was divided by reported weights to derive MET equivalents (1 MET = 1 kcal/kg/hour). If unavailable, body weight was calculated from reported BMI and height. If BMI was reported without height, a mean value from national survey data was used for height.

#### 2.4.4 Subtracting resting energy expenditure from estimates of PA

To marginalize studies reporting volume of PA in gross units, 1 MET-h was subtracted for each hour of activity. If mean duration was not available, we used a conversion equation derived from all remaining studies where both volume and duration were available (Supplement, eMethods 5).

#### 2.4.5 Isolating the non-occupational component of aggregate PA estimates

Some articles provided aggregated exposures of non-occupational and occupational PA alongside other behaviors, such as sleep or sedentary time. When quantified information about these behaviors was available, this was subtracted from the point estimate of each exposure category. If no quantifiable data were available, we assumed occupational activity to be 40 hours/week at 1.25 MET (or 0.25 mMET) or the value of the lowest exposure category and subtracted this from all exposure categories.

### 2.5 Meta-analytic methods

We conducted meta-analysis for any exposure-outcome pair with at least four independent results. Where necessary, risk estimates were re-calculated to set the least active category as the referent.^14^ For studies reporting only stratified results (e.g., sex or ethnicity), we combined stratum-specific risks into overall population estimates using fixed-effect meta-analysis.

We performed a two-stage random-effects meta-analysis. In the first stage, we estimated the study-specific associations using generalized least squares to incorporate the correlation within each set of log-relative risks.^15, 16^ In the second stage, we estimated the pooled association by combining study-specific dose-response coefficients using restricted maximum likelihood.^17, 18^ We assumed non-linearity of dose-response associations,^4, 5, 7, 19-21^ and thus modelled them by fitting restricted cubic splines. Given that the volume of PA reported in most studies were at the lower end of the exposure range and that there is greater uncertainty about reliability of very high levels of self-reported PA, we set three knots at the 0^th^, 37.5^th^, and 75^th^ percentiles of person-years rather than persons (0^th^, 42.5^th^ and 85^th^ percentiles in sensitivity analyses). The slope was fixed at the last knot. If the statistical model was unable to converge, we progressively increased the percentile for the upper knot by one percent until model convergence.

To investigate the potential effect of study-level confounders on the pooled results across outcomes, we conducted subgroup analyses with the 11 studies that reported results for all all-cause, CVD, and cancer mortality outcomes and contrasted them with results from the corresponding main analysis. We also conducted subgroup analysis by sex using studies that reported separate results for men and women.

### 2.6 Estimation of population health impact

Potential population impact fractions (PIFs) were calculated for all outcomes based on PA exposure levels in the population of all included cohorts for a given outcome.^22^ PIFs were calculated for three exposure levels based on the World Health Organization PA recommendations for adults^23^: 8.75 mMET-h/week (the minimum recommended level, equivalent to 2.5 hours/week of PA at an intensity of 3.5 mMET, such as a brisk walking), 17.5 mMET-h/week (upper bound of recommended levels for health benefits), and 4.375 mMET-h/week (half the minimum recommended level).

### 2.7 Risk of bias assessment

We explored the impact of four potential sources of bias in individual articles and our meta-analytical procedures: how studies had analyzed participants with other morbidities, whether they excluded early incident cases during follow-up, whether imputation procedures for missing data were required, and how we handled exposure harmonization including separation of occupational PA. For each of these we contrasted the overall risk estimates between studies with different characteristics. This was done for the five outcomes with the largest number of independent results (all-cause mortality, and total CVD and cancer mortality and incidence), using a fixed PA level of 8.75 mMET-h/week, relevant to the four sources of bias.

### 2.8 Software, data, and code availability

Analyses were performed using R, version 4.0.5, and the dosresmeta package,^18^ version 2.0.1. An interactive interface to visualize the dose-response associations was developed using the Shiny package, version 1.0.5. Syntax for all analyses and the interactive interface are available at *https://github.com/meta-analyses/*.

### 2.9 Patient and public involvement

Patients and the public were not involved in the development of this work.

## 3. RESULTS

### 3.1 Identified literature

A total of 196 articles^7, 24-218^ were included (Supplement, eFigure 1), covering 94 cohorts and 330 independent results. The evidence base was largest for all-cause mortality (50 independent results; 163,415,543 person-years, 811,616 events), total CVD incidence (37 independent results; 28,884,209 person-years, 74,757 events), and total cancer incidence (31 independent results; 35,500,867 person-years, 185,870 events). Details of all selected articles can be found in our <u>OSF repository</u> and data underlying each of the dose-response estimation can be downloaded at *https://shiny.mrc-epid.cam.ac.uk/meta-analyses-physical-activity/*.

### 3.2 Primary dose-response analyses

Most study participants reported non-occupational PA levels below 17.5 mMET-h/week (76% of person-years), with almost all data below 35 mMET-h/week (94% of person-years). Figure 1 shows the exposure distribution for the cohorts included in the all-cause mortality analysis. Inverse, curvilinear dose-response associations between PA and most outcomes were observed, with stronger associations at lower volumes of PA. In most cases, diminishing reductions in risk and increasing uncertainty were observed at high PA volumes, particularly beyond 17.5 mMET-h/week. Interactive dose-response curves and the exposure distributions are available at https://shiny.mrc-epid.cam.ac.uk/meta-analyses-physical-activity/.

**Figure 1.**
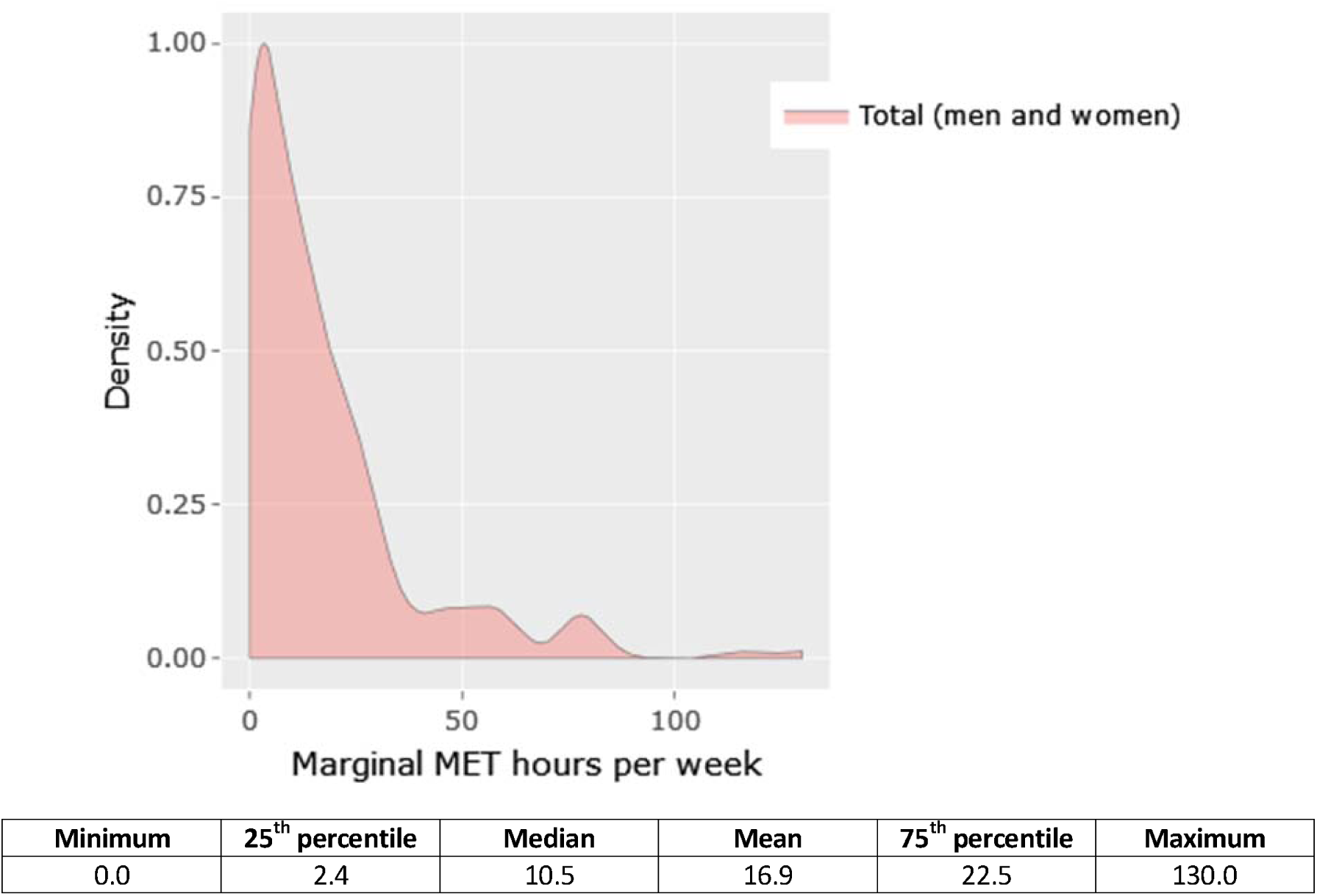
Distribution of marginal MET hours per week for cohorts included in the all-cause mortality analysis. Exposure distribution for cohorts included in the analysis of other outcomes are available at https://shiny.mrc-epid.cam.ac.uk/meta-analyses-physical-activity/.

The association was stronger and more curvilinear for all-cause and CVD mortality than for cancer mortality (Figure 2, Table 1). Compared to inactive individuals, adults accumulating 8.75 mMET-h/week had 31% (95%CI: 27 to 35%) and 29% (95%CI: 23 to 34%) lower risk of all-cause and CVD mortality, respectively, whereas the risk reduction for total cancer mortality was 15% (95%CI: 11 to 19%).

**Figure 2.**
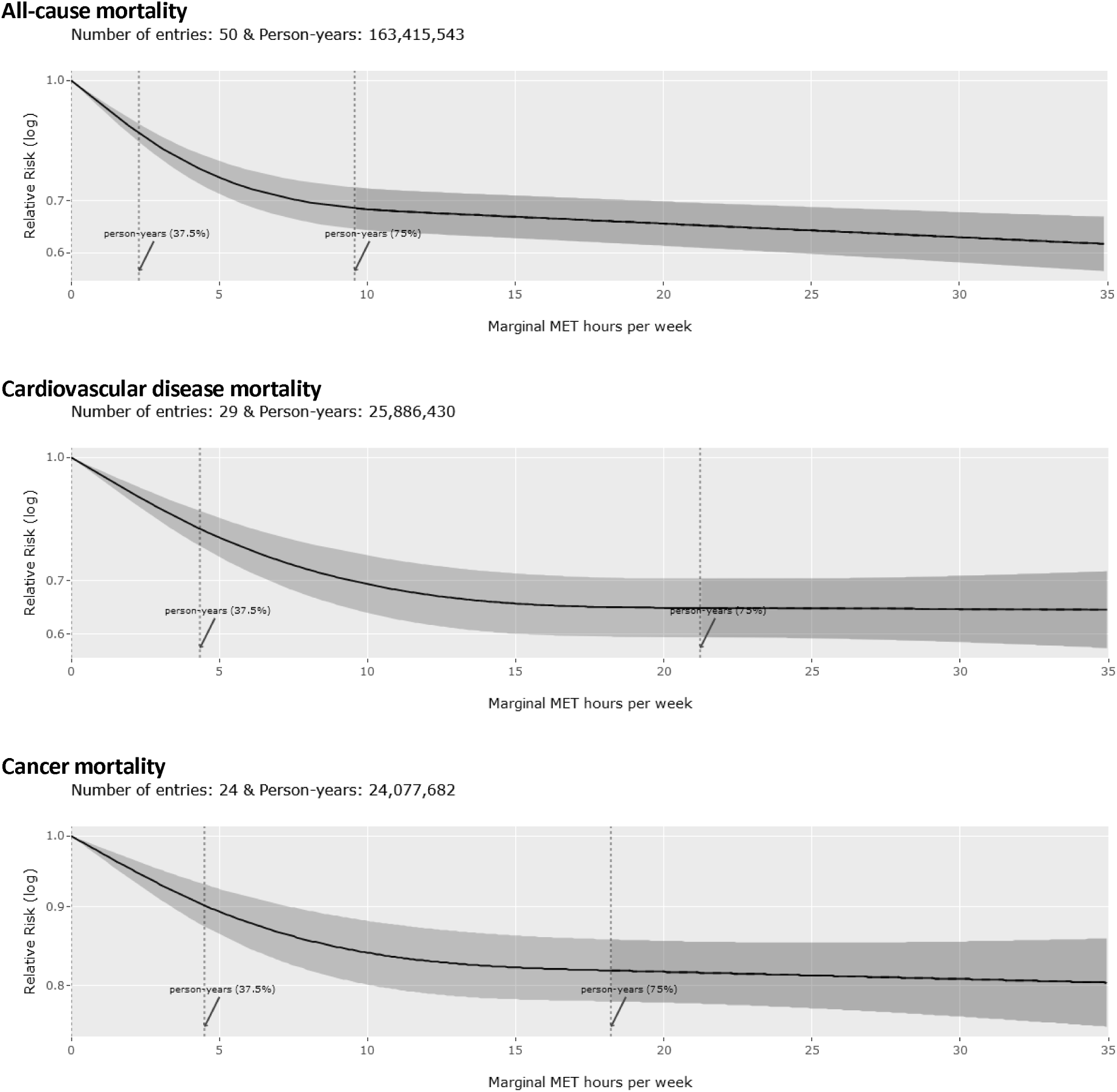
Dose-response association between non-occupational physical activity and mortality outcomes. Vertical dashed lines: cubic spline knots (0^th^, 37.5^th^, and 75^th^ percentile of person-years) Dark grey area beyond upper knot: constrained to be linear.

**Table 1.** Relative risk of mortality and incidence of cardiovascular diseases and cancers at three physical activity levels, and respective potential impact fractions.

| Outcomes | 4.375 mMET-h/week |  | 8.75 mMET-h/week |  | 17.5 mMET-h/week |  |
| --- | --- | --- | --- | --- | --- | --- |
|  | RR (95% CI) | PIF (95% CI) | RR (95% CI) | PIF (95% CI) | RR (95% CI) | PIF (95% CI) |
| **Mortality** |  |  |  |  |  |  |
| All-cause mortality | 0.77<br>(0.73 - 0.80) | 10.11<br>(8.32 - 11.91) | 0.69<br>(0.65 - 0.73) | 15.66<br>(13.16 - 18.15) | 0.66<br>(0.62 - 0.70) | 18.75<br>(16.09 - 21.39) |
| Total CVD | 0.81<br>(0.77 - 0.85) | 6.02<br>(4.42 - 7.69) | 0.71<br>(0.66 - 0.77) | 12.25<br>(9.37 - 15.17) | 0.65<br>(0.60 - 0.71) | 17.05<br>(13.92 - 20.19) |
| Total cancer | 0.90<br>(0.88 - 0.93) | 3.68<br>(2.54 - 4.86) | 0.85<br>(0.81 - 0.89) | 7.05<br>(5.1 - 9.00) | 0.82<br>(0.78 - 0.86) | 9.29<br>(7.36 - 11.22) |
| **CVD incidence** |  |  |  |  |  |  |
| Total CVD | 0.83<br>(0.79 - 0.87) | 5.29<br>(3.93 - 6.70) | 0.73<br>(0.69 - 0.79) | 10.86<br>(8.43 - 13.31) | 0.67<br>(0.63 - 0.72) | 15.42<br>(13.01 - 17.85) |
| Coronary heart disease | 0.86<br>(0.83 - 0.90) | 5.51<br>(3.95 - 7.11) | 0.79<br>(0.74 - 0.84) | 10.28<br>(7.61 - 12.96) | 0.74<br>(0.69 - 0.80) | 13.58<br>(10.51 - 16.64) |
| Stroke | 0.86<br>(0.83 - 0.90) | 5.53<br>(3.96 - 7.15) | 0.80<br>(0.75 - 0.85) | 9.77<br>(7.31 - 12.24) | 0.77<br>(0.72 - 0.82) | 12.37<br>(9.63 - 15.10) |
| Heart failure | 0.90<br>(0.85 - 0.96) | 2.23<br>(0.80 - 3.89) | 0.80<br>(0.75 - 0.93) | 4.63<br>(1.90 - 7.66) | 0.77<br>(0.68 - 0.87) | 7.86<br>(4.31 - 11.68) |
| **Cancer incidence** |  |  |  |  |  |  |
| Total cancer | 0.93<br>(0.91 - 0.95) | 2.53<br>(1.66 - 3.42) | 0.88<br>(0.85 - 0.92) | 5.22<br>(3.55 - 6.90) | 0.85<br>(0.81 - 0.89) | 7.80<br>(5.65 - 9.93) |
| Head and neck | 0.74<br>(0.59 - 0.94) | 10.59<br>(2.10 - 19.96) | 0.65<br>(0.46 - 0.91) | 17.46<br>(3.68 - 30.98) | 0.63<br>(0.42 - 0.94) | 19.83<br>(1.23 - 36.55) |
| Myeloid leukaemia | 0.80<br>(0.66 - 0.97) | 4.93<br>(0.68 - 9.70) | 0.75<br>(0.60 - 0.95) | 7.08<br>(1.65 - 12.94) | 0.75<br>(0.60 - 0.94) | 7.57<br>(2.23 - 13.33) |
| Myeloma | 0.84<br>(0.74 - 0.95) | 3.34<br>(0.90 - 6.26) | 0.75<br>(0.61 - 0.92) | 6.76<br>(1.90 - 12.09) | 0.72<br>(0.57 - 0.92) | 8.30<br>(1.91 - 14.99) |
| Gastric cardia | 0.86<br>(0.78 - 0.95) | 1.95<br>(0.58 - 3.68) | 0.78<br>(0.66 - 0.91) | 4.77<br>(1.58 - 8.57) | 0.73<br>(0.60 - 0.88) | 6.77<br>(2.55 - 11.63) |
| Lung | 0.89<br>(0.87 - 0.92) | 4.39<br>(3.16 - 5.63) | 0.84<br>(0.81 - 0.88) | 7.26<br>(5.10 - 9.40) | 0.83<br>(0.76 - 0.91) | 8.54<br>(3.04 - 13.98) |
| Liver | 0.90<br>(0.83 - 0.98) | 3.49<br>(0.58 - 6.71) | 0.84<br>(0.73 - 0.96) | 6.75<br>(1.44 - 12.31) | 0.78<br>(0.66 - 0.93) | 10.27<br>(3.26 - 17.31) |
| Endometrial | 0.94<br>(0.90 - 0.99) | 1.10<br>(0.26 - 2.02) | 0.90<br>(0.84 - 0.97) | 2.62<br>(0.76 - 4.60) | 0.87<br>(0.80 - 0.95) | 4.54<br>(1.88 - 7.28) |
| Colon | 0.96<br>(0.93 - 0.99) | 0.70<br>(0.11 - 1.33) | 0.93<br>(0.87 - 0.99) | 1.69<br>(0.33 - 3.12) | 0.90<br>(0.84 - 0.97) | 2.96<br>(0.92 - 5.05) |
| Breast | 0.97<br>(0.96 - 0.99) | 0.69<br>(0.32 - 1.07) | 0.95<br>(0.92 - 0.97) | 1.62<br>(0.79 - 2.46) | 0.92<br>(0.88 - 0.96) | 3.23<br>(1.85 - 4.63) |
| Bladder | 0.93<br>(0.84 - 1.02) | 1.91<br>(-0.54 - 4.52) | 0.90<br>(0.80 - 1.02) | 2.98<br>(-0.24 - 6.33) | 0.90<br>(0.81 - 1.01) | 3.11<br>(0.02 - 6.35) |
| Rectum | 0.96<br>(0.92 - 1.01) | 0.77<br>(-0.15 - 1.75) | 0.95<br>(0.88 - 1.02) | 1.43<br>(-0.45 - 3.36) | 0.96<br>(0.88 - 1.04) | 0.52<br>(-1.74 - 2.82) |
| Esophageal | 0.97<br>(0.89 - 1.05) | 0.68<br>(-0.95 - 2.68) | 0.95<br>(0.82 - 1.09) | 1.38<br>(-2.06 - 5.32) | 0.94<br>(0.79 - 1.12) | 1.57<br>(-3.05 - 6.67) |
| Prostate | 1.00<br>(0.99 - 1.02) | -0.10<br>(-0.48 - 0.29) | 1.01<br>(0.98 - 1.04) | -0.21<br>(-1.10 - 0.71) | 1.01<br>(0.96 - 1.05) | -0.10<br>(-1.61 - 1.42) |
| Kidney | 1.02<br>(0.92 - 1.13) | -0.65<br>(-3.69 - 2.81) | 1.03<br>(0.89 - 1.19) | -1.23<br>(-6.39 - 4.34) | 1.04<br>(0.88 - 1.24) | -1.91<br>(-9.20 - 5.63) |

mMET = marginal metabolic equivalent of task; RR = relative risk; PIF = potential impact fraction; 95% CI = 95% confidence interval. CVD = cardiovascular disease.

A strong curvilinear association was observed for total CVD incidence (27% lower risk [95%CI: 21 to 31%] at 8.75 mMET-h/week). However, associations were weaker and more linear for the incidence of specific CVD outcomes (coronary heart disease, heart failure, and stroke), with the strongest association observed for coronary heart disease (21% lower risk [95%CI: 16 to 26%] at 8.75 mMET-h/week) (Figure 3, Table 1).

**Figure 3.**
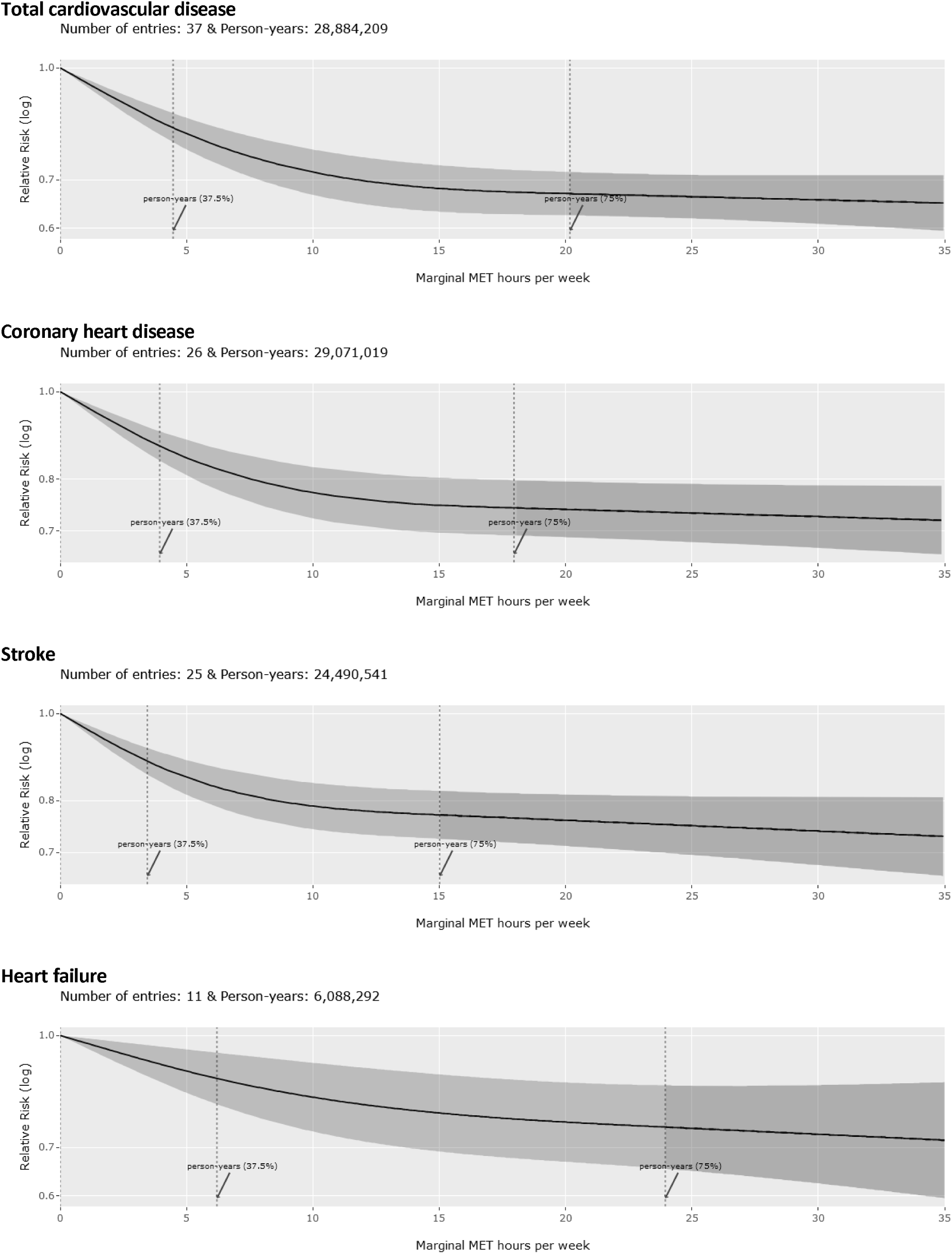
Dose-response association between non-occupational physical activity and incidence of cardiovascular diseases. Vertical dashed lines: cubic spline knots (0^th^, 37.5^th^, and 75^th^ percentile of person-years) Dark grey area beyond upper knot: constrained to be linear.

The association was weaker and more linear for total cancer incidence (12% lower risk [95%CI: 8 to 15%] at 8.75 mMET-h/week). For the incidence of site-specific cancers, curvilinear and stronger associations were observed for head and neck, myeloid leukemia, myeloma, and gastric cardia (35 to 22% lower risk at 8.75 mMET-h/week). Weaker and more linear associations were observed for lung, liver, endometrial, colon, and breast (16 to 5% lower risk at 8.75 mMET-h/week). Non-significant associations were observed for bladder, esophageal, prostate, and rectal cancer (Figure 4, Table 1). No eligible studies were available for malignant melanoma.

**Figure 4.**
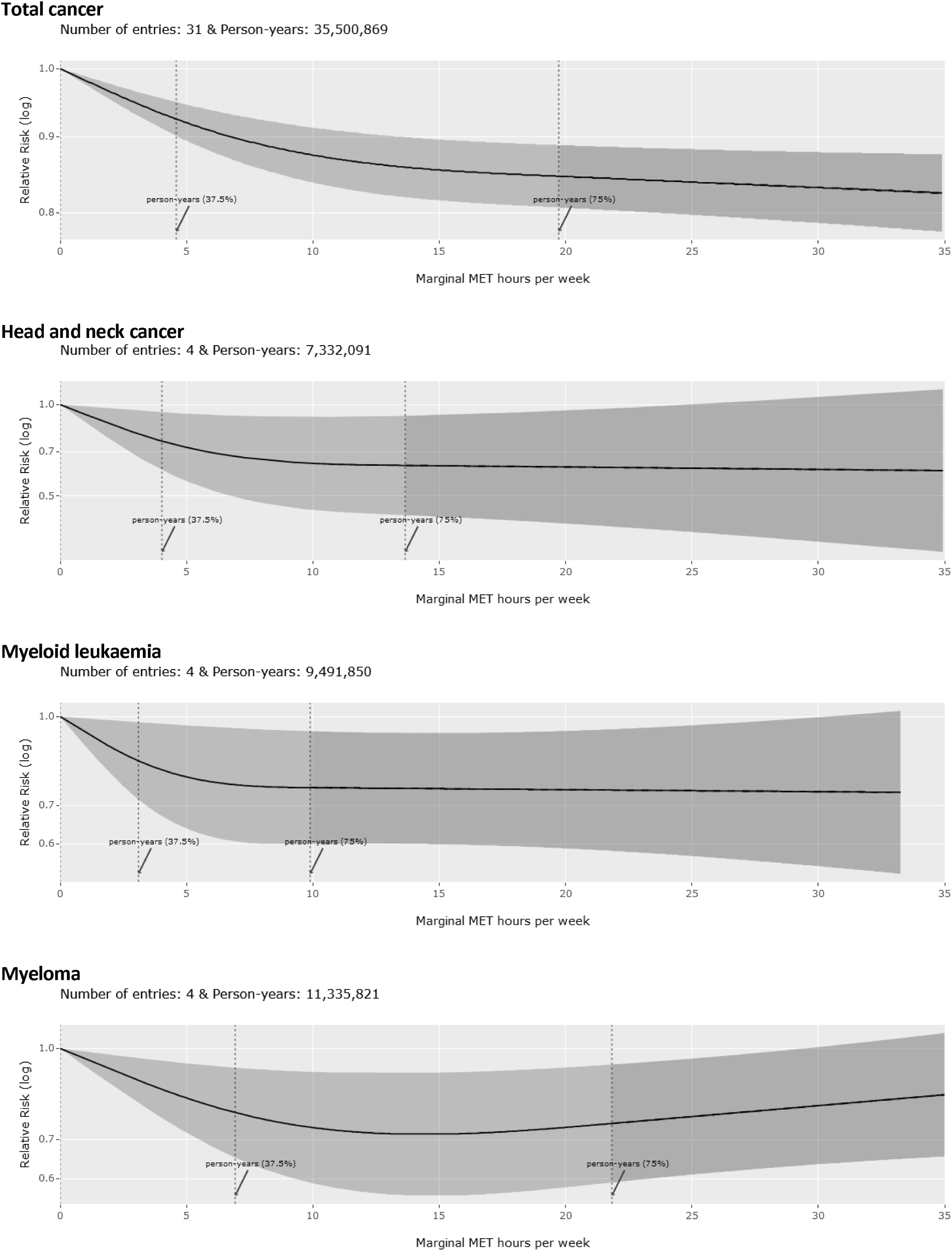
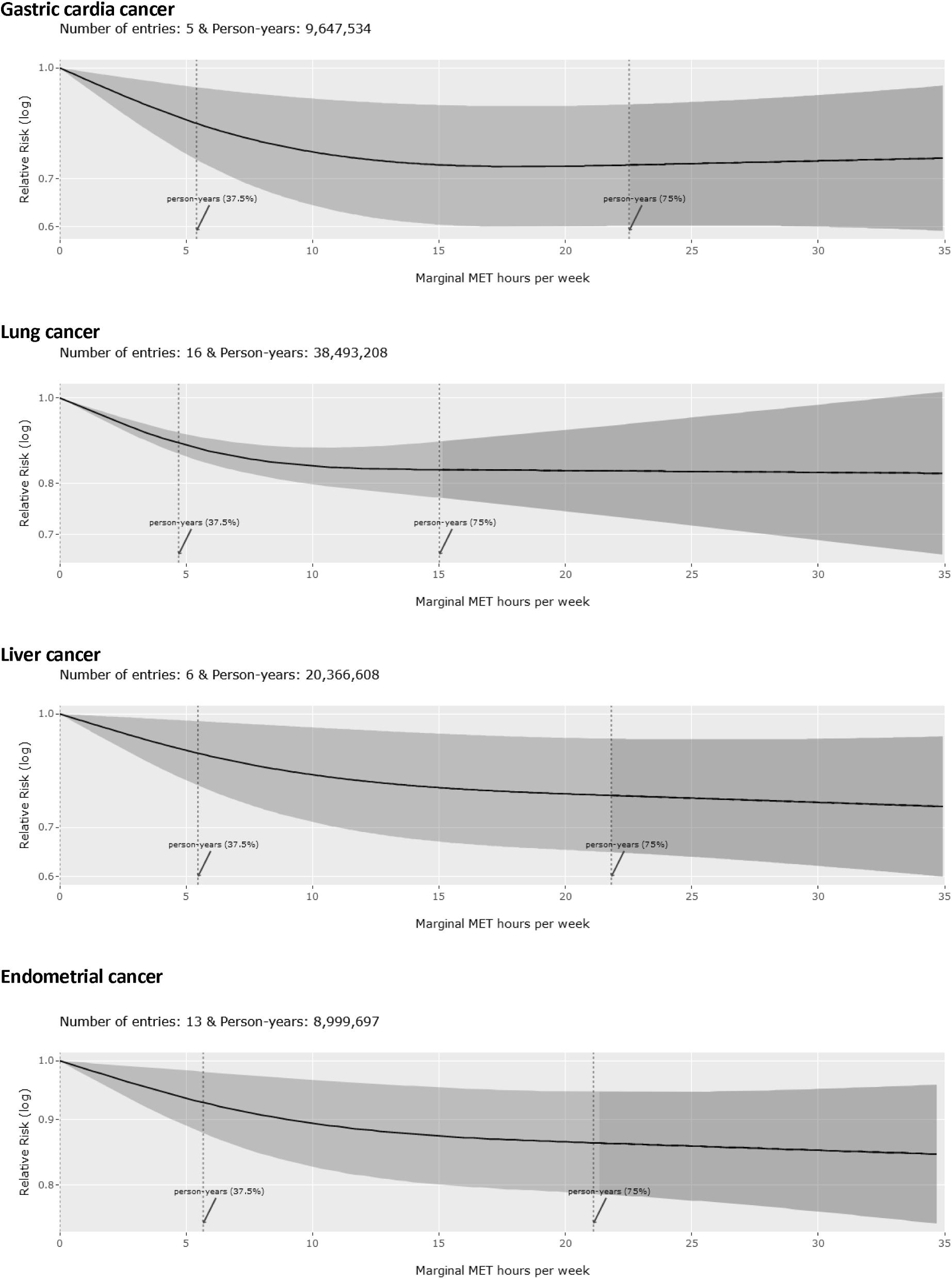
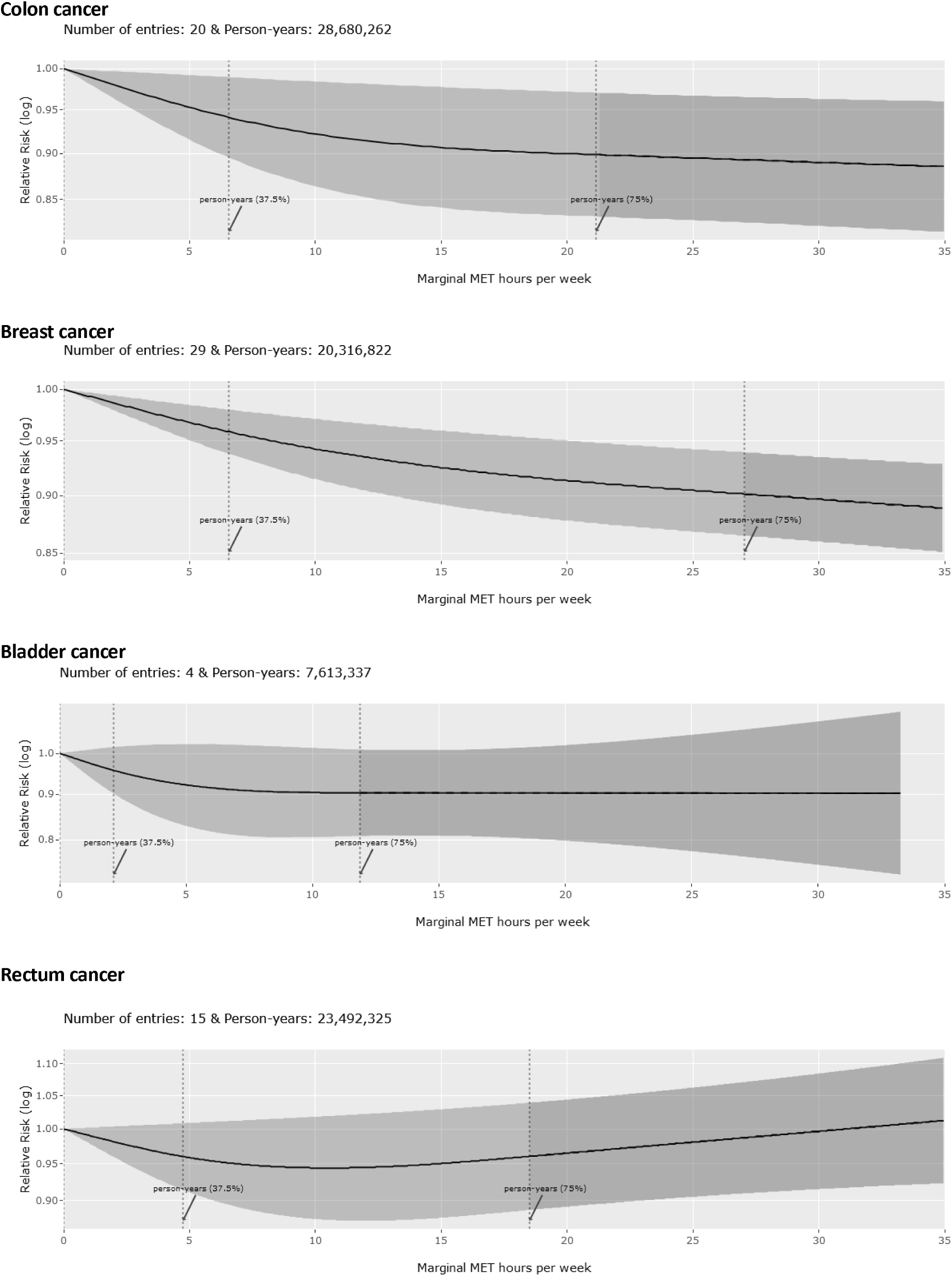
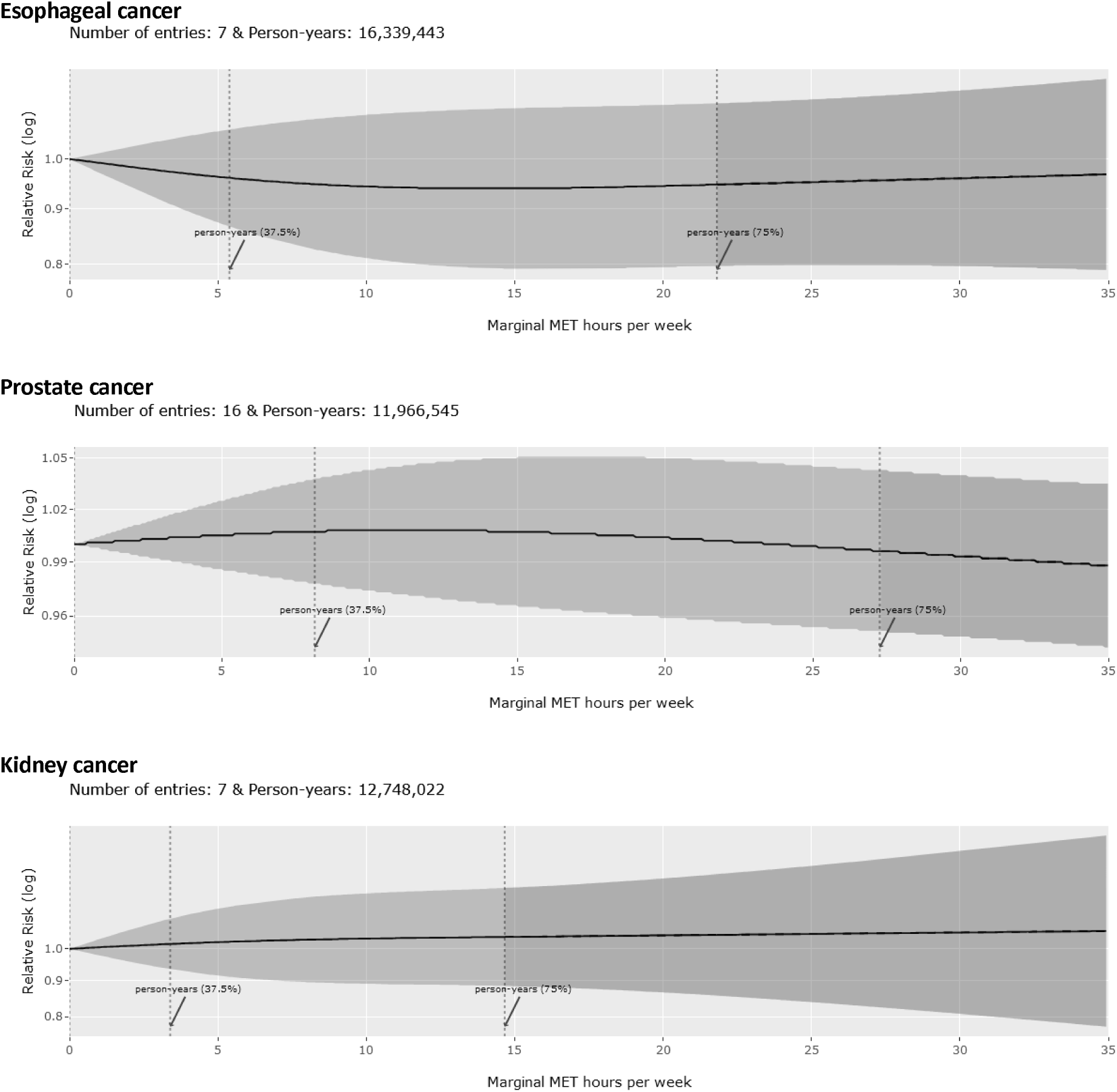
Dose-response association between non-occupational physical activity and incidence of cancers. Vertical dashed lines: cubic spline knots (0^th^, 37.5^th^, and 75^th^ percentile of person-years) Dark grey area beyond upper knot: constrained to be linear.

### 3.3 Potential population impact

Assuming a causal relationship between non-occupational PA and the examined health outcomes, shifting the distribution of non-occupational PA in the cohorts toward the recommended levels would have averted a substantial fraction of the population disease burden. For example, if all individuals accumulated at least the recommended level of PA (8.75 mMET-h/week), then 15.7% (95%CI: 13.1 to 18.2%), 12.3% (95%CI: 9.4 to 15.2%), and 7.1% (95%CI: 5.1 to 9.0%) of all-cause, CVD-, and cancer-related deaths, respectively, would potentially have been averted. Additionally, 10.9% (95%CI: 8.4 to 13.3%) and 5.2% (95%CI: 3.6 to 6.9%) of all incident cases of CVD and cancer would have been prevented. Notably, 10.1% (95%CI: 8.3 to 11.9%) of all deaths would have been prevented if all adults achieved at least half the recommended PA level (Table 1).

### 3.4 Subgroup analysis

#### 3.4.1 Pooled estimates from studies that reported results for all-cause, CVD, and cancer mortality

The pooled analysis from the 11 studies that reported results for all-cause, CVD, and cancer mortality showed weaker associations for all-cause mortality and stronger associations for CVD and cancer mortality (Supplement, eResults 1).

#### 3.4.2 Sex-stratified results

Sex-stratified results were available for all-cause, CVD, and cancer mortality and incidence of total CVD, coronary heart disease, stroke, total cancer, and cancers of the colon and rectum. There was some evidence of stronger associations in women than in men for all-cause and CVD mortality and CVD incidence, but the reverse for cancer mortality and incidence (Supplement, eResults 2).

### 3.5 Risk of bias

Across the five outcomes with the largest number of independent results (all-cause mortality, and total CVD and cancer mortality and incidence), 83 to 93% of the studies used statistical adjustments to control for other morbidities, often combined with exclusion of adults with some more serious pre-existing conditions at baseline. Studies that predominantly used statistical adjustments tended to report stronger associations for CVD mortality compared with studies that excluded adults with pre-existing conditions at baseline. Between 54 and 59% of the studies did not exclude mortality and disease events occurring within the first few years of follow-up, and those that did tended to report weaker associations for all-cause mortality and CVD mortality and incidence. Data imputation was required for 58 to 64% of the studies, but no evidence of bias was found. The most frequently used procedures for exposure harmonization were measurement unit conversions (42 to 49% of studies) and the assumption of standard values for the intensity and duration of PA reported (39 to 50%), but no evidence of bias was found. In 75% of the studies, the original exposure assessment did not include occupational PA and no evidence of bias was observed for studies requiring domain separation (Supplement, eResults 3).

### 3.6 Sensitivity analyses

In studies that did not report the duration or intensity of PA, assuming (where appropriate) shorter session duration (0.5 instead of 0.75 hour) and lower intensity (1 mMET lower for moderate and vigorous PA) did not materially alter the dose-response associations for most outcomes, especially analyses which included the largest number of independent results (Supplement, eResults 4). When shorter duration and lower intensity were used, associations were stronger for CVD outcomes, particularly at lower PA volumes. Placing the restricted cubic spline knots at the 42.5^th^ and 85^th^ percentiles resulted in less stable estimations at higher PA volumes, with those specific segments of the dose-response curves being based on less data and with a higher risk of exposure misclassification.

Sensitivity analyses using analytical models that did not include adjustment for adiposity showed stronger associations for all-cause and CVD mortality, and the incidence of total CVD, heart failure, and site-specific cancers of the endometrium, kidney, lung, and rectum (Supplement, eResults 4). Notably, the association was significant for rectal cancer.

## 4. DISCUSSION

We present comprehensive dose-response meta-analyses of the associations between non-occupational PA and a wide range of mortality and disease outcomes. Our extensive exposure data harmonization allowed inclusion of a larger evidence base than previous meta-analyses for 17/22 health outcomes investigated, including associations for nine site-specific cancers that have not previously been reported.

We showed inverse non-linear dose-response associations for all-cause mortality and a range of CVD and cancer outcomes. This suggests that the greatest potential benefits can be achieved through small increases in non-occupational PA from an inactive lifestyle, with incrementally smaller additional benefits up to approximately 17.5 mMET-h/week. Above this level, the evidence base was weaker. Shifting population levels of activity toward achieving the minimum PA recommendations of 8.75 mMET-h/week (equivalent to 150 min/week of moderate-to-vigorous PA) would potentially have prevented 16% of all premature deaths recorded.

Compared with previous studies, we found similar associations for all-cause^7, 20^ and cancer mortality,^21^ and stronger associations and narrower confidence intervals for CVD mortality^219, 220^ at the recommended levels of PA. For cancers of the bladder, breast, colon, endometrium, esophagus, liver and rectum, our results were largely similar to that of a recent pooled analysis.^19^ However, we found stronger associations for cancers of the gastric cardia, head and neck, myeloid leukemia, and myeloma, whereas the association for kidney cancer was non-significant. Contrasting with Moore *et al’*s^11^ finding of harmful effects of PA on prostate cancer, our results did not show an association. Comparisons should be interpreted with caution as most previous studies have focused solely on leisure-time PA^7, 11, 19, 20, 220^ and meta-analyses by Li *et al*^21^ and Wahid *et al*^219^ included studies that assessed PA in any domain.

Compared with results from the GBD,^5^ which have assessed total PA rather than non-occupational PA, we found stronger associations for heart disease and stroke at the minimum recommended PA level. Associations reported by the GBD at approximately 130 MET-h/week for total PA were observed in our study at 17.5 mMET-h/week for non-occupational PA, a much lower volume even considering that we excluded energy expenditure from the resting metabolic rate and occupational activity. Given the challenges of assessing occupational activity, estimates of total PA from self-report are often implausibly high.^221^ Hence, the importance of PA may have been underestimated by the GBD. This, combined with the uncertainties around the health benefits of occupational PA,^8, 9^ means our results may be more relevant from a public health perspective.

It is surprising that the risk reduction for all-cause mortality (31% at minimum recommended PA level) is similar to that for total CVD (29%) and much larger than for total cancer mortality (15%). Equally, it is surprising that the association for total CVD incidence is stronger than that for coronary heart disease or stroke. Weaker associations would typically be expected for more composite outcomes (e.g., all-cause mortality and total CVD incidence) than for the outcomes within them for which a significant association has been established. Potential explanations include detection bias for some disease events, misclassification of causes of death (especially for more specific disease outcomes), differential residual confounding, reverse causality by outcome, and different inclusion criteria by disease outcome, such as the inclusion of different disease groups which may have stronger associations with PA.^222^ There may also be study-level confounders, and this is supported by our post-hoc subgroup analysis, which showed that studies reporting all three mortality outcomes found stronger associations for CVD than for all-cause mortality. Taken together, it is probable that the risk estimates for all-cause mortality are more accurate than for specific disease outcomes.

A strength of our study was the use of comprehensive exposure data harmonization methods that enabled the inclusion of a larger evidence base. Another strength is the use of more sophisticated meta-analytic methods that allows the shape of the dose-response association to vary across the exposure range, rather than linear models of transformed exposure or grouping doses within high versus low PA or multiple exposure intervals. We improved the placement of spline knots compared to our previous work^4^ by considering the distribution of person-years instead of non-weighted associations across the PA range, acknowledging the paucity of data at higher volumes of PA.

Our study also has limitations. Many of the included studies relied on self-reported questionnaires without any validation or calibration. In the exposure data harmonization procedures, it was also sometimes necessary to make assumptions about parameters of physical activity such as intensity and duration where these were not explicitly reported. Nonetheless, our sensitivity analyses demonstrated the robustness of our results to the assumptions made. Measurement error is likely to lead to an underestimation of the true association between PA and the various outcomes, as demonstrated by the stronger associations seen in studies using device-based measures of PA.^223^ Misclassification of some outcomes may be possible as procedures for the ascertainment of outcomes may not be consistent across all studies. Residual confounding and reverse causation could remain. To mitigate confounding, the most comprehensively adjusted risk estimates were used. However, the level of covariate adjustment and the degree to which these covariates effectively control for confounding may not be consistent across all studies. We excluded studies with less than three years of follow up to mitigate reverse causation.^224^ Although the bias from reverse causation diminishes with longer periods of follow-up, changes in PA level over time could be possible.^225, 226^

Our findings support the PA recommendations of 150-300 minutes/week of moderate PA (or 75-150 minutes/week of vigorous PA, or an equivalent combination of moderate and vigorous activities),^23^ in that this exposure level generally seems to equate to maximum or near to maximal benefits. However, the dose-response associations also demonstrate that appreciable health benefits can be gained from 75 minutes/week or less of moderate activity (i.e., half the recommended minimum level of activity). Thus, our findings support the recent change in messaging to “doing some PA is better than doing none”, and suggest that the emphasis on threshold-based recommendations could be further reduced.

It should be noted that our exposure estimates are derived from a variety of self-reported questionnaires that capture mostly moderate-to-vigorous activities. These exposure estimates differ from those derived using device-based measures, which also record light intensity and intermittent activities that are more difficult to recall.^223, 227, 228^ Self-report and device-based measures are therefore complimentary but not interchangeable,^228^ an important consideration when formulating public health messages.

Future research should investigate reasons for the apparent inconsistencies in dose-response associations between composite and individual disease endpoints. Although our risk of bias and sensitivity analyses showed robustness to the approaches we took during data handling and dose harmonization, future studies could quantify methodological uncertainties (e.g., inaccuracies in exposure assessment) and propagate them in the etiological analyses to provide more realistic uncertainty ranges for the dose-response associations. The evidence base was weaker for higher volumes of activity and further research is required to ascertain the shape of associations more reliably at the higher end of the PA continuum.

## 5. CONCLUSION

We found evidence of dose-dependent associations between increasing non-occupational PA and a wide range of mortality, CVD, and cancer outcomes. Results showed stronger associations for all-cause and CVD mortality, and weaker associations for the incidence of cancer including variation by site. Appreciable population health benefits can be gained from 75 minutes/week or less of moderate non-occupational PA activity (i.e., half the recommended minimum level of activity).

## Supporting information

MOOSE checklist

## Data Availability

All data produced are available online at https://shiny.mrc-epid.cam.ac.uk/meta-analyses-physical-activity/

https://osf.io/pnvdc/?view_only=d7d8338ed63a486c8038ead389f34159

https://shiny.mrc-epid.cam.ac.uk/meta-analyses-physical-activity/

## Authors’ contributions

*Conception and design*: Garcia, Pearce, Mok, Strain, Woodcock, Brage.

*Data acquisition*: Garcia, Pearce, Mok, Strain, Ali, Dempsey, Golubic, Kelly, Laird, McNamara, Moore, Sa, Smith, Wijndaele.

*Statistical analysis*: Garcia, Pearce, Abbas, Crippa, Woodcock, Brage.

*Software*: Abbas.

*Interpretation of data*: Garcia, Pearce, Mok, Strain, Ali, Crippa, Dempsey, Golubic, Kelly, Laird, McNamara, Moore, Sa, Smith, Wijndaele, Woodcock, Brage.

*Drafting of the manuscript*: Garcia, Pearce, Mok, Strain, Sa, Woodcock, Brage.

*Critical revision of the manuscript*: All authors.

All authors approved the submitted version and agreed to be accountable for all aspects of the work.

## Guarantors

Garcia, Pearce, Abbas, Mok, Strain, Woodcock, and Brage had full access to the data, accept full responsibility for the work, and controlled the decision to publish. The corresponding author (Brage) attests that all listed authors meet authorship criteria and that no others meeting the criteria have been omitted.

## Additional contributions

We thank the authors of original articles who replied to our request for additional information about their analyses: Prof Alejandro Arrieta, Dr Michael Cook, Prof Ulf Ekelund, Prof Edward Giovannucci, Prof Reza Malekzadeh, Dr Jessica Morris, Dr Neil Murphy, Dr Mahdi Nalini, Dr Nicole Niehoff, Dr Tom Nilsen, Dr Annlia Paganini-Hill, Dr Song-Yi Park, Prof Louise Russell, and Prof Wei Zheng. We also thank the IT team of the MRC Epidemiology Unit (University of Cambridge) for setting up the server to host the online results.

