## Supplement for "Non-occupational physical activity and risk of 22 cardiovascular disease, cancer, and mortality outcomes: a dose-response meta-analysis of large prospective studies"

### **eMethods 1: study inclusion and exclusion criteria**

| **Include** | **Exclude** |
| --- | --- |
| **Participants** | |
| Human adults (18+ years of age) | Cohorts of institutionalised adults, medical patients (i.e., in secondary or tertiary prevention) or high-risk populations (e.g., hypertension, diabetes - except conditions inevitable with ageing, as post-menopause) |
| **Exposure** | |
| Volume (marginal MET-h/week) of non-occupational physical activity – this includes leisure-time physical activity alone, combined with other non-occupational physical activity domains (e.g., transport and/or domestic activity), or combined with specific types of activity such as walking. Physical activity volume can be either self-reported or measured by devices and must be reported in at least three exposure levels | Measures of physical activity from which occupational activity cannot be factored out; individual domains of physical activity which occur outside of work but do not include leisure-time activity (e.g., travel physical activity only); other risk factors either alone or combined with physical activity |
| **Comparators** | |
| Lower volumes of physical activity (as defined in ‘Interventions’) |  |
| **Outcomes** | |
| 1) All-cause mortality 2) Cardiovascular diseases (fatal and non-fatal):    a. Total CVD (I00-I99)*    b. Stroke (I60-I66)    c. Heart failure (I50, I11.0, I13.0, I13.2)    d. Coronary heart disease (I20-I25) 3) Cancers** (fatal and non-fatal):    a. Total cancer (C00-D49)    b. Breast (C50)    c. Endometrial (C54)    d. Lung (C34)    e. Colon (C18)    f. Esophageal (C15)    g. Liver (C22-C24)    h. Kidney (C64-C65)    i. Gastric cardia (C16.0)    j. Myeloid leukemia (C92)    k. Myeloma (C90)    l. Head and neck (C76.0)    m. Rectum (C20)    n. Bladder (C67)    o. Prostate (C61)    p. Malignant melanoma (C43)   Cases ascertained via health, death, cancer, hospital or insurance registers; death certificates; participants and their next-of-kin | Less than three years of follow-up period |
| **Study design** | |
| General population prospective cohort studies (including case-cohort and nested case-control studies) with sample size larger than 10,000 participants; results must be reported either as hazard ratios, relative risks or odds ratios with confidence intervals | Other study designs (e.g., experimental or cross-sectional), even when nested in prospective cohort studies |
| * Diseases coded using the International statistical classification of diseases and related health problems, 10th revision.^1^  ** Cancers associated with leisure-time physical activity in Moore et al.^2^ | |

- Type of documents: peer-reviewed articles published in academic journals.
- No limits on language and date of publication.

### **eMethods 2: search strings**

**Pubmed**

((physical activit*[Title/Abstract]) OR (physical inactivity[Title/Abstract]) OR (exercise[Title/Abstract]) OR (exercises[Title/Abstract]) OR (exercising[Title/Abstract]) OR (recreational activit*[Title/Abstract]) OR (sport[Title/Abstract]) OR (sports[Title/Abstract]) OR (active transport[Title/Abstract]) OR (active transportation[Title/Abstract]) OR (active commut*[Title/Abstract]) OR (active travel*[Title/Abstract]) OR (household activit*[Title/Abstract]) OR (housework[Title/Abstract]) OR (non-exercise activit*[Title/Abstract]) OR (nonexercise activit*[Title/Abstract]) OR (activities of daily living[Title/Abstract]) OR (bicycle[Title/Abstract]) OR (bicycling[Title/Abstract]) OR (bike[Title/Abstract]) OR (biking[Title/Abstract]) OR (walk[Title/Abstract]) OR (walking[Title/Abstract]) OR (run[Title/Abstract]) OR (running[Title/Abstract]) OR (jogging[Title/Abstract]) OR (swim[Title/Abstract]) OR (swimming[Title/Abstract]) OR (resistance training[Title/Abstract]) OR (weight lifting[Title/Abstract]) OR (physical fitness[Title/Abstract]) OR (physical endurance[Title/Abstract]) OR (activity energy expenditure[Title/Abstract]) OR (caloric expenditure[Title/Abstract]) OR (motor activity[Mesh:NoExp]) OR (exercise[Mesh:NoExp]) OR (human physical conditioning[Mesh]) OR (leisure activities[Mesh:NoExp]) OR (recreation[Mesh:NoExp]) OR (dancing[Mesh]) OR (gardening[Mesh]) OR (sports[Mesh]) OR (activities of daily living[Mesh:NoExp]))

AND

((mortality[Title/Abstract]) OR (cardiovascular disease*[Title/Abstract]) OR (acute coronary syndrome[Title/Abstract]) OR (myocardial infarction[Title/Abstract]) OR (myocardial ischemia[Title/Abstract]) OR (heart disease*[Title/Abstract]) OR (coronary disease*[Title/Abstract]) OR (coronary artery disease*[Title/Abstract]) OR (heart failure[Title/Abstract]) OR (cardiac failure[Title/Abstract]) OR (stroke[Title/Abstract]) OR (brain ischemia[Title/Abstract]) OR (cerebrovascular accident*[Title/Abstract]) OR (cerebrovascular disorder*[Title/Abstract]) OR (cerebrovascular disease*[Title/Abstract]) OR (intracranial arterial disease*[Title/Abstract]) OR (ganglia hemorrhage[Title/Abstract]) OR (ganglia haemorrhage[Title/Abstract]) OR (cerebral intraventricular hemorrhage[Title/Abstract]) OR (cerebral intraventricular haemorrhage[Title/Abstract]) OR (hypertensive intracranial hemorrhage[Title/Abstract]) OR (hypertensive intracranial haemorrhage[Title/Abstract]) OR (cerebral haemorrhage[Title/Abstract]) OR (cerebral hemorrhage[Title/Abstract]) OR (cancer[Title/Abstract]) OR (cancers[Title/Abstract]) OR (tumour[Title/Abstract]) OR (tumours[Title/Abstract]) OR (tumor[Title/Abstract]) OR (tumors[Title/Abstract]) OR (neoplasm[Title/Abstract]) OR (neoplasms[Title/Abstract]) OR (mortality[Mesh:NoExp]) OR (cause of death[Mesh]) OR (premature mortality[Mesh]) OR (cardiovascular diseases[Mesh:NoExp]) OR (heart diseases[Mesh:NoExp]) OR (myocardial ischemia[Mesh]) OR (heart failure[Mesh]) OR (cerebrovascular disorders[Mesh:NoExp]) OR (brain ischemia[Mesh]) OR (cerebral small vessel diseases[Mesh]) OR (intracranial arterial diseases[Mesh]) OR (basal ganglia hemorrhage [Mesh]) OR (cerebral intraventricular hemorrhage [Mesh]) OR (hypertensive intracranial hemorrhage [Mesh]) OR (stroke[Mesh]) OR (neoplasms[Mesh]))

AND

((cohort[Title/Abstract]) OR (cohorts[Title/Abstract]) OR (follow-up study[Title/Abstract]) OR (follow-up studies[Title/Abstract]) OR (prospective study[Title/Abstract]) OR (prospective studies[Title/Abstract]) OR (longitudinal study[Title/Abstract]) OR (longitudinal studies[Title/Abstract]) OR (cohort studies[Mesh:NoExp]) OR (follow-up studies[Mesh]) OR (longitudinal studies[Mesh:NoExp]) OR (prospective studies[Mesh]))

**Scopus**

(TITLE-ABS-KEY(“physical activit*”) OR TITLE-ABS-KEY(“physical inactivity”) OR TITLE-ABS-KEY(exercise) OR TITLE-ABS-KEY(exercises) OR TITLE-ABS-KEY(exercising) OR TITLE-ABS-KEY(“recreational activit*”) OR TITLE-ABS-KEY(sport) OR TITLE-ABS-KEY(sports) OR TITLE-ABS-KEY(“active transport”) OR TITLE-ABS-KEY(“active transportation”) OR TITLE-ABS-KEY(“active commut*”) OR TITLE-ABS-KEY(“active travel*”) OR TITLE-ABS-KEY(“household activit*”) OR TITLE-ABS-KEY(housework) OR TITLE-ABS-KEY(“non-exercise activit*”) OR TITLE-ABS-KEY(“nonexercise activit*”) OR TITLE-ABS-KEY(“activities of daily living”) OR TITLE-ABS-KEY(bicycle) OR TITLE-ABS-KEY(bicycling) OR TITLE-ABS-KEY(bike) OR TITLE-ABS-KEY(biking) OR TITLE-ABS-KEY(walk) OR TITLE-ABS-KEY(walking) OR TITLE-ABS-KEY(run) OR TITLE-ABS-KEY(running) OR TITLE-ABS-KEY(jogging) OR TITLE-ABS-KEY(swim) OR TITLE-ABS-KEY(swimming) OR TITLE-ABS-KEY(“resistance training”) OR TITLE-ABS-KEY(“weight lifting”) OR TITLE-ABS-KEY(“physical fitness”) OR TITLE-ABS-KEY(“physical endurance”) OR TITLE-ABS-KEY(“activity energy expenditure”) OR TITLE-ABS-KEY(“caloric expenditure”))

AND

(TITLE-ABS-KEY(mortality) OR TITLE-ABS-KEY(“cardiovascular disease*”) OR TITLE-ABS-KEY(“acute coronary syndrome”) OR TITLE-ABS-KEY(“myocardial infarction”) OR TITLE-ABS-KEY(“myocardial ischemia”) OR TITLE-ABS-KEY(“heart disease*”) OR TITLE-ABS-KEY(“coronary disease*”) OR TITLE-ABS-KEY(“coronary artery disease*”) OR TITLE-ABS-KEY(“heart failure”) OR TITLE-ABS-KEY(“cardiac failure”) OR TITLE-ABS-KEY(stroke) OR TITLE-ABS-KEY(“brain ischemia”) OR TITLE-ABS-KEY(“cerebrovascular accident*”) OR TITLE-ABS-KEY(“cerebrovascular disorder*”) OR TITLE-ABS-KEY(“cerebrovascular disease*”) OR TITLE-ABS-KEY(“intracranial arterial disease*”) OR TITLE-ABS-KEY(“ganglia hemorrhage") OR TITLE-ABS-KEY(“ganglia haemorrhage”) OR TITLE-ABS-KEY(“cerebral intraventricular hemorrhage”) OR TITLE-ABS-KEY(“cerebral intraventricular haemorrhage”) OR TITLE-ABS-KEY(“hypertensive intracranial hemorrhage") OR TITLE-ABS-KEY(“hypertensive intracranial haemorrhage") OR TITLE-ABS-KEY(“cerebral hemorrhage”) OR TITLE-ABS-KEY(“cerebral haemorrhage”) OR TITLE-ABS-KEY(cancer) OR TITLE-ABS-KEY(cancers) OR TITLE-ABS-KEY(tumour) OR TITLE-ABS-KEY(tumours) OR TITLE-ABS-KEY(tumor) OR TITLE-ABS-KEY(tumors) OR TITLE-ABS-KEY(neoplasm) OR TITLE-ABS-KEY(neoplasms))

AND

(TITLE-ABS-KEY(cohort) OR TITLE-ABS-KEY(cohorts) OR TITLE-ABS-KEY(“follow-up study”) OR TITLE-ABS-KEY(“follow-up studies”) OR TITLE-ABS-KEY(“prospective study”) OR TITLE-ABS-KEY(“prospective studies”) OR TITLE-ABS-KEY(“longitudinal study”) OR TITLE-ABS-KEY(“longitudinal studies”))

**Web of Science**

(TS=(“physical activit*”) OR TS=(“physical inactivity”) OR TS=(exercise) OR TS=(exercises) OR TS=(exercising) OR TS=(“recreational activit*”) OR TS=(sport) OR TS=(sports) OR TS=(“active transport”) OR TS=(“active transportation”) OR TS=(“active commut*”) OR TS=(“active travel*”) OR TS=(“household activit*”) OR TS=(housework) OR TS=(“non-exercise activit*”) OR TS=(“nonexercise activit*”) OR TS=(“activities of daily living”) OR TS=(bicycle) OR TS=(bicycling) OR TS=(bike) OR TS=(biking) OR TS=(walk) OR TS=(walking) OR TS=(run) OR TS=(running) OR TS=(jogging) OR TS=(swim) OR TS=(swimming) OR TS=(“resistance training”) OR TS=(“weight lifting”) OR TS=(“physical fitness”) OR TS=(“physical endurance”) OR TS=(“activity energy expenditure”) OR TS=(“caloric expenditure”))

AND

(TS=(mortality) OR TS=(“cardiovascular disease*”) OR TS=(“acute coronary syndrome”) OR TS=(“myocardial infarction”) OR TS=(“myocardial ischemia”) OR TS=(“heart disease*”) OR TS=(“coronary disease*”) OR TS=(“coronary artery disease*”) OR TS=(“heart failure”) OR TS=(“cardiac failure”) OR TS=(stroke) OR TS=(“brain ischemia”) OR TS=(“cerebrovascular accident*”) OR TS=(“cerebrovascular disorder*”) OR TS=(“cerebrovascular disease*”) OR TS=(“intracranial arterial disease*”) OR TS=(“ganglia hemorrhage") OR TS=(“ganglia haemorrhage”) OR TS=(“cerebral intraventricular hemorrhage”) OR TS=(“cerebral intraventricular haemorrhage”) OR TS=(“hypertensive intracranial hemorrhage") OR TS=(“hypertensive intracranial haemorrhage") OR TS=(“cerebral hemorrhage”) OR TS=(“cerebral haemorrhage”) OR TS=(cancer) OR TS=(cancers) OR TS=(tumour) OR TS=(tumours) OR TS=(tumor) OR TS=(tumors) OR TS=(neoplasm) OR TS=(neoplasms))

AND

(TS=(cohort) OR TS=(cohorts) OR TS=(“follow-up study”) OR TS=(“follow-up studies”) OR TS=(“prospective study”) OR TS=(“prospective studies”) OR TS=(“longitudinal study”) OR TS=(“longitudinal studies”))

### **eMethods 3: article selection criteria when multiple reports on the same cohort and outcome were available**

The following criteria were used in the indicated hierarchy. That is, if two or more article were found and they balanced each other out in terms of the criteria (i.e., one was better for one criterion, but the other one for a second criterion), this hierarchy was then used to decide which article to select.

1) Lower risk of bias (exposure assessment, outcome ascertainment, exclusion of cases at baseline, exclusion of participants with relevant comorbidities at baseline or inclusion of baseline comorbidities as covariates in the regression model, and exclusion of new cases in the first years of follow-up).
2) Preferred measure of physical activity (e.g., MET-h/day rather than sessions).
3) More physical activity levels.
4) Separate results for men and women.
5) Longer follow-up period.

### **eMethods 4: data imputation procedures**

**CRITICAL INFORMATION (required to run the meta-analysis)**

**Number of individuals in each exposure category (required only when risk estimate presented as relative risk or odds ratio):**

1) Estimated via proportional weighting using the following equation:

| $\frac{p_{i}}{P}S$ | (1) |
| --- | --- |

Where *p_i_* is person-years in the *i*^th^ exposure category, *P* is total person-years, and *S* is total analytical sample size.

2) Or estimated via proportional weighting using the following steps (when person-years in each exposure category was not reported):

$$w_{i}=\frac{c_{i}}{\left( c_{1}{RR}_{i} \right)}$$

(2)

$$s_{1}=\frac{S}{\sum_{i=1}^{n} w_{i}}$$

(3)

$$s_{i}=w_{i}s_{1}$$

(4)

Where *w_i_* is the ratio between person-years in the *i*^th^ exposure category and in the reference exposure category, *c_i_* is cases in the *i*^th^ exposure category (subscript 1 refers to the reference level), *RR_i_* is minimally adjusted relative risk in the *i*^th^ exposure category, *s_i_* is number of individuals in the *i*^th^ exposure category, and *S* is total analytical sample size.

Equations 3 and 5 derive from:

$${RR}_{i}=\frac{\frac{c_{i}}{s_{i}}}{\frac{c_{1}}{s_{1}}}=\frac{c_{i}s_{1}}{c_{1}s_{i}} \to s_{i}=\frac{c_{i}s_{1}}{c_{1}{RR}_{i}} \to\frac{s_{i}}{s_{1}}=\frac{c_{i}}{c_{1}{RR}_{i}}$$

(5)

Equation 4 derives from:

$$S=\sum_{i=1}^{n} s_{i}=\sum_{i=1}^{n} w_{i}s_{1}$$

(6)

Once $s_{1}$is found, it can be used in Equation 5 to estimate number of individuals in all other exposure categories.

3) Or authors contacted.

**Person-years in each exposure category (required only when risk estimate presented as hazard ratio):**

1) Estimated via proportional weighting using the following equation:

| $\frac{n_{i}}{S}P$ | (7) |
| --- | --- |

Where *n_i_* is number of individuals in the *i*^th^ exposure category, *S* is total analytical sample size, and *P* is total person-years.

2) Or estimated via proportional weighting (when number of individuals in each exposure category was not reported) using Equations 3 to 5 substituting the number of individuals in the *i*^th^ exposure category (*s_i_*) and total analytical sample size (*S*) by person-years in the *i*^th^ exposure category and total person-years, respectively.
3) Or authors contacted.

**Cases in each exposure category:**

1) Estimated via proportional weighting using the following steps:

$$c_{1_{t}}=\frac{{RR}_{1}}{\sum_{i=1}^{n} {RR}_{i}}C$$

(8)

$$r_{1}=\frac{c_{1_{t}}}{n_{1}}$$

(9)

$$r_{i}={RR}_{i}r_{1}$$

(10)

$$c_{i}=n_{i}r_{i}\frac{C}{\sum_{i=1}^{n} (n_{i}r_{i})}$$

(11)

Where *c_i_* is cases in the *i*^th^ exposure category (subscript 1 refers to the reference level; subscript *t* indicates temporary (non-corrected) number of cases), *RR_i_* is risk estimate in the *i*^th^ exposure category, *C* is total number of cases, *r_i_* is risk in the *i*^th^ exposure category, *n_i_* is person-years (or number of individuals) in the *i*^th^ exposure category. Equation 11 is also used to estimate the final number of cases in the reference level.

2) Or authors contacted.

**BACKGROUND INFORMATION (obtained or estimated only when needed to impute at least one of the critical information)**

**Total analytical sample size:**

1) Calculated via summation of the number of individuals in each exposure category.
2) Or reported in the article.
3) Or estimated dividing total person-years by mean follow-up.
4) Or authors contacted.

**Mean follow-up:**

1) Reported in the article (mean or median follow-up).
2) Or calculated by dividing total person-years by total analytical sample size.
3) Or reported study follow-up duration (e.g., 1990 to 2010 = 20 years), as an approximate surrogate.
4) Or authors contacted.

**Total person-years:**

1) Calculated via summation of person-years in each exposure category.
2) Or reported in the article.
3) Or estimated multiplying total analytical sample size by mean follow-up.
4) Or authors contacted.

**Total number of cases:**

1) Calculated via summation of cases in each exposure category.
2) Or reported in the article.
3) Or authors contacted.

### **eMethods 5: estimating the resting component of energy expenditure to convert MET to mMET**

Total physical activity volume can be calculated by multiplying duration by the rate of energy expenditure (intensity). Rate of energy expenditure is often expressed in gross units of metabolic equivalent of task (MET), which includes both a resting (1 MET) and an activity component (the remainder, e.g., 3.5 METs for a 4.5 gross MET activity).

When publications report activity volume in total MET-hours (hours * MET) per week, it is necessary to obtain information on the duration of activities making up this total such that 1 MET can be subtracted for each hour to remove the resting component and give physical activity energy expenditure in marginalised MET (mMET).

In the example below, a study reports total physical activity volume as 9 MET-hours. Knowledge of the duration of activity of two hours is necessary to calculate and remove the resting component giving marginalised MET (mMET).

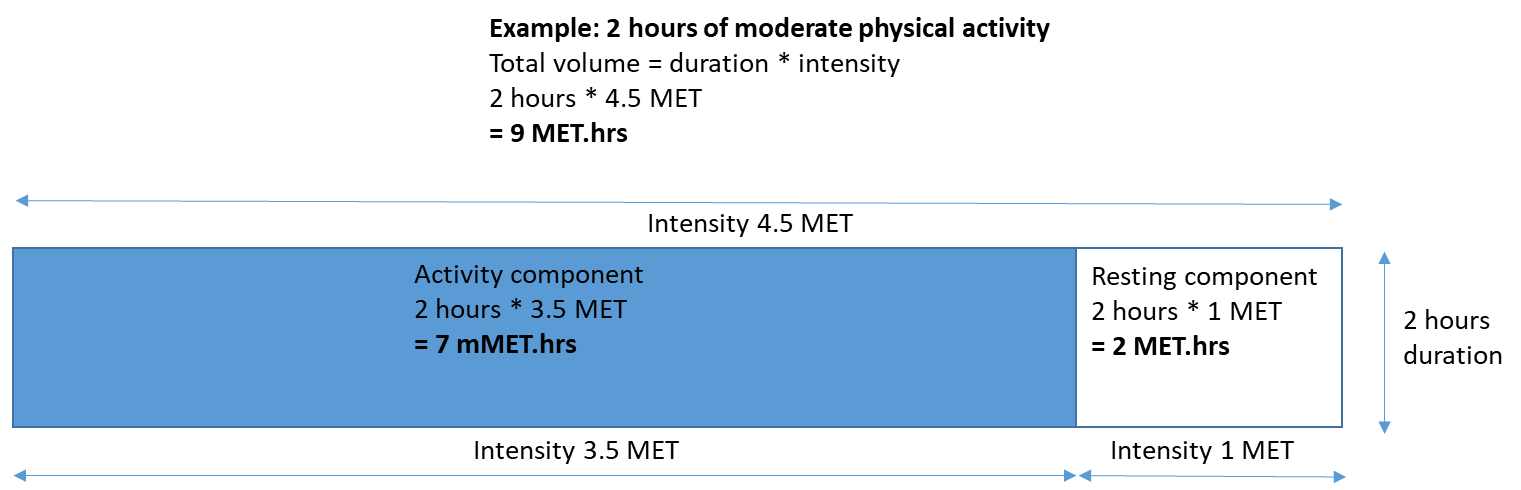

For studies that did not provide activity duration, a conversion equation from gross to marginal units is required. For the current analyses, we used a regression equation derived from the studies that provided both estimates, weighted for sample size per exposure category:

$$mMEThours=0.89*METhours-0.52*{METhours}^{0.5}$$

(1)

The studies/cohorts contributing to this equation were:

| **Authors and year** | **Cohort** | **Participants** |
| --- | --- | --- |
| Leet et al.^3^ | Aerobics Center Longitudinal Study | Men and women aged 20-82 years |
| Chomistek et al.^4^ | Health Professionals Follow-up Study | Men aged 40-75 years |
| Chomistek et al.^5^* | Women's Genome Health Study | Women aged 45+ years |
| Wen et al.^6^ | Taiwan Medical Screening Program | Men and women aged 20+ years |
| Mok et al.^7^ | Severance Cohort Study | Men and women aged 30-93 years |
| Hu et al.^8^ | Nurses’ Health Study | Women aged 40-65 years |
| Besson et al.^9^* | EPIC-Norfolk | Men and women aged 40-79 years |
| * Identified by systematic literature review and used in equation derivation but not included in main analyses because full results were available in a parent study. | | |

### **eFigure 1: study screening and selection flowchart**

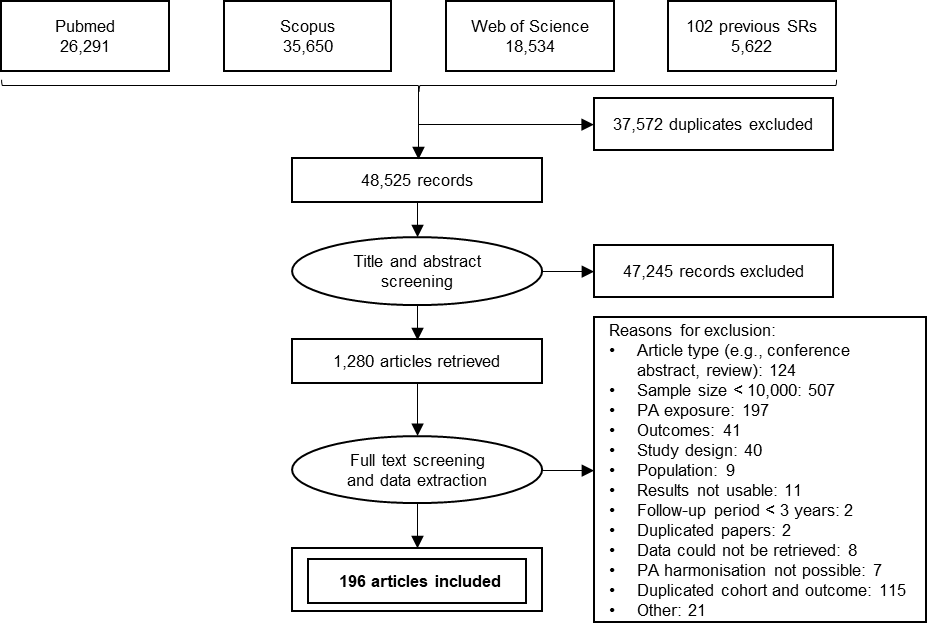

SR: systematic reviews. PA: physical activity.

A list of articles not included in the analysis and the reason for exclusion can be obtained with the authors.

### **eResults 1: meta-analysis of the 11 studies that reported all-cause, cardiovascular and cancer mortality risk**

| **Outcomes** | **4.375 mMET-h/week** | **8.75 mMET-h/week** | **17.5 mMET-h/week** |
| --- | --- | --- | --- |
|  | **RR (95% CI)** | **RR (95% CI)** | **RR (95% CI)** |
| Subset of 11 studies^6, 10-19^ |  |  |  |
| All-cause mortality | 0.81  (0.74 - 0.89) | 0.67  (0.57 - 0.78) | 0.61  (0.52 - 0.73) |
| Total CVD | 0.77  (0.69 - 0.85) | 0.67  (0.57 - 0.78) | 0.61  (0.52 - 0.73) |
| Total cancer | 0.89  (0.85 - 0.94) | 0.84  (0.79 - 0.90) | 0.81  (0.76 - 0.86) |
| All studies |  |  |  |
| All-cause mortality | 0.77  (0.73 - 0.80) | 0.69  (0.65 - 0.73) | 0.66  (0.62 - 0.70) |
| Total CVD | 0.81  (0.77 - 0.85) | 0.71  (0.66 - 0.77) | 0.65  (0.60 - 0.71) |
| Total cancer | 0.90  (0.88 - 0.93) | 0.85  (0.81 - 0.89) | 0.82  (0.78 - 0.86) |

### **eResults 2: sex-stratified results**

| **Relative risk of mortality and incidence of cardiovascular diseases and cancers at three physical activity levels, by sex.** | | | | | | | | |
| --- | --- | --- | --- | --- | --- | --- | --- | --- |
| **Outcomes** | **4.375 mMET-h/week** | |  | **8.75 mMET-h/week** | |  | **17.5 mMET-h/week** | |
|  | **Men** | **Women** |  | **Men** | **Women** |  | **Men** | **Women** |
|  | **RR (95% CI)** | **RR (95% CI)** |  | **RR (95% CI)** | **RR (95% CI)** |  | **RR (95% CI)** | **RR (95% CI)** |
| Mortality |  |  |  |  |  |  |  |  |
| All-cause mortality | 0.88  (0.85 - 0.91) | 0.84  (0.8 - 0.87) |  | 0.80  (0.76 - 0.85) | 0.71  (0.66 - 0.76) |  | 0.75  (0.71 - 0.80) | 0.71  (0.66 - 0.76) |
| Total CVD | 0.87  (0.82 - 0.94) | 0.81  (0.74 - 0.88) |  | 0.80  (0.72 - 0.89) | 0.67  (0.60 - 0.77) |  | 0.75  (0.68 - 0.82) | 0.67  (0.60 - 0.77) |
| Total cancer | 0.90  (0.87 - 0.94) | 0.95  (0.92 - 0.98) |  | 0.86  (0.80 - 0.91) | 0.88  (0.83 - 0.93) |  | 0.84  (0.79 - 0.09) | 0.88  (0.83 - 0.93) |
| CVD incidence |  |  |  |  |  |  |  |  |
| Total CVD | 0.89  (0.84 - 0.93) | 0.84  (0.76 - 0.91) |  | 0.81  (0.75 - 0.88) | 0.70  (0.62 - 0.79) |  | 0.76  (0.71 - 0.82) | 0.70  (0.62 - 0.79) |
| Coronary heart disease | 0.92  (0.85 - 0.99) | 0.84  (0.75 - 0.93) |  | 0.88  (0.79 - 0.98) | 0.73  (0.60 - 0.87) |  | 0.86  (0.76 - 0.97) | 0.73  (0.60 - 0.87) |
| Stroke | 0.90  (0.86 - 0.94) | 0.88  (0.79 - 0.97) |  | 0.85  (0.79 - 0.91) | 0.76  (0.66 - 0.88) |  | 0.82  (0.76 - 0.89) | 0.76  (0.66 - 0.88) |
| Cancer incidence |  |  |  |  |  |  |  |  |
| Total cancer | 0.95  (0.92 - 0.98) | 0.98  (0.96 - 0.99) |  | 0.92  (0.86 - 0.97) | 0.93  (0.89 - 0.97) |  | 0.89  (0.82 - 0.96) | 0.93  (0.89 - 0.97) |
| Colon | 0.91  (0.86 - 0.96) | 0.97  (0.92 - 1.02) |  | 0.85  (0.78 - 0.94) | 0.93  (0.84 - 1.03) |  | 0.81  (0.72 - 0.91) | 0.93  (0.84 - 1.03) |
| Rectum | 0.92  (0.80 - 1.07) | 1.09  (0.90 - 1.33) |  | 0.90  (0.74 - 1.1) | 1.13  (0.73 - 1.76) |  | 0.92  (0.76 - 1.11) | 1.13  (0.73 - 1.76) |
| mMET = marginal metabolic equivalent of task; RR = relative risk; 95% CI = 95% confidence interval. CVD = cardiovascular disease. | | | | | | | | |

| **Potential impact fraction of physical activity on mortality and incidence of cardiovascular diseases and cancers at three physical activity levels, by sex.** | | | | | | | | |
| --- | --- | --- | --- | --- | --- | --- | --- | --- |
| **Outcomes** | **4.375 mMET-h/week** | |  | **8.75 mMET-h/week** | |  | **17.5 mMET-h/week** | |
|  | **Men** | **Women** |  | **Men** | **Women** |  | **Men** | **Women** |
|  | **PIF (95% CI)** | **PIF (95% CI)** |  | **PIF (95% CI)** | **PIF (95% CI)** |  | **PIF (95% CI)** | **PIF (95% CI)** |
| Mortality |  |  |  |  |  |  |  |  |
| All-cause mortality | 3.74  (2.69 - 4.83) | 5.43  (4.00 - 6.94) |  | 7.39  (5.52 - 9.31) | 9.82  (7.42 - 12.29) |  | 10.69  (8.37 - 13.03) | 12.30  (9.30 - 15.35) |
| Total CVD | 3.16  (1.51 - 4.86) | 6.15  (3.54 - 8.85) |  | 7.78  (4.48 - 11.02) | 12.83  (8.06 - 17.52) |  | 11.82  (9.62 - 14.04) | 15.65  (11.65 - 19.64) |
| Total cancer | 2.72  (1.57 - 3.90) | 1.56  (0.67 - 2.48) |  | 5.13  (3.11 - 7.18) | 3.51  (1.68 - 5.36) |  | 6.21  (4.20 - 8.25) | 6.27  (3.94 - 8.59) |
| CVD incidence |  |  |  |  |  |  |  |  |
| Total CVD | 2.78  (1.54 - 4.05) | 4.95  (2.45 - 7.54) |  | 6.64  (4.17 - 9.10) | 10.4  (5.90 - 14.84) |  | 10.06  (8.23 - 11.91) | 13.65  (10.48 - 16.86) |
| Coronary heart disease | 2.99  (0.38 - 5.72) | 7.10  (2.81 - 11.68) |  | 5.07  (0.74 - 9.44) | 12.17  (4.98 - 19.42) |  | 6.15  (1.21 - 11.06) | 14.60  (6.11 - 22.94) |
| Stroke | 3.21  (1.72 - 4.80) | 4.71  (1.02 - 8.60) |  | 6.12  (3.47 - 8.89) | 8.90  (2.92 - 14.91) |  | 7.93  (4.87 - 11.07) | 12.79  (7.22 - 18.38) |
| Cancer incidence |  |  |  |  |  |  |  |  |
| Total cancer | 1.38  (0.40 - 2.41) | 0.73  (0.28 - 1.19) |  | 3.09  (0.94 - 5.29) | 1.77  (0.72 - 2.84) |  | 4.94  (1.63 - 8.21) | 3.92  (1.81 - 6.04) |
| Colon | 2.07  (0.78 - 3.48) | 0.67  (-0.44 - 1.87) |  | 4.13  (1.68 - 6.71) | 1.39  (-0.80 - 3.68) |  | 6.42  (3.07 - 9.85) | 2.25  (-0.55 - 5.11) |
| Rectum | 1.90  (-1.52 - 5.71) | -2.64  (-7.02 - 3.40) |  | 3.10  (-2.79 - 9.21) | -4.66  (-10.03 - 2.64) |  | 1.43  (-3.83 - 6.99) | -4.57  (-31.13 - 20.26) |
| mMET = marginal metabolic equivalent of task; PIF = potential impact fraction; 95% CI = 95% confidence interval. CVD = cardiovascular disease. | | | | | | | | |

### **eResults 3: risk of bias assessment**

For the risk of bias assessment, we classified the harmonisation methods in two ways. Firstly, we classified studies depending on whether we made assumptions about the occupational component of an aggregated PA exposure (occupational PA not included in exposure; or assumptions about occupational PA made). Secondly, we described the overall method of converting exposure data to mMET-h/week (measurement unit conversion from MET, kcal, or kJ data; calibration using validation studies; using assumptions on ≤ 2 of intensity, duration, or frequency; or conversion of non-numeric data using assumptions on intensity, duration, and frequency).

**All-cause mortality**

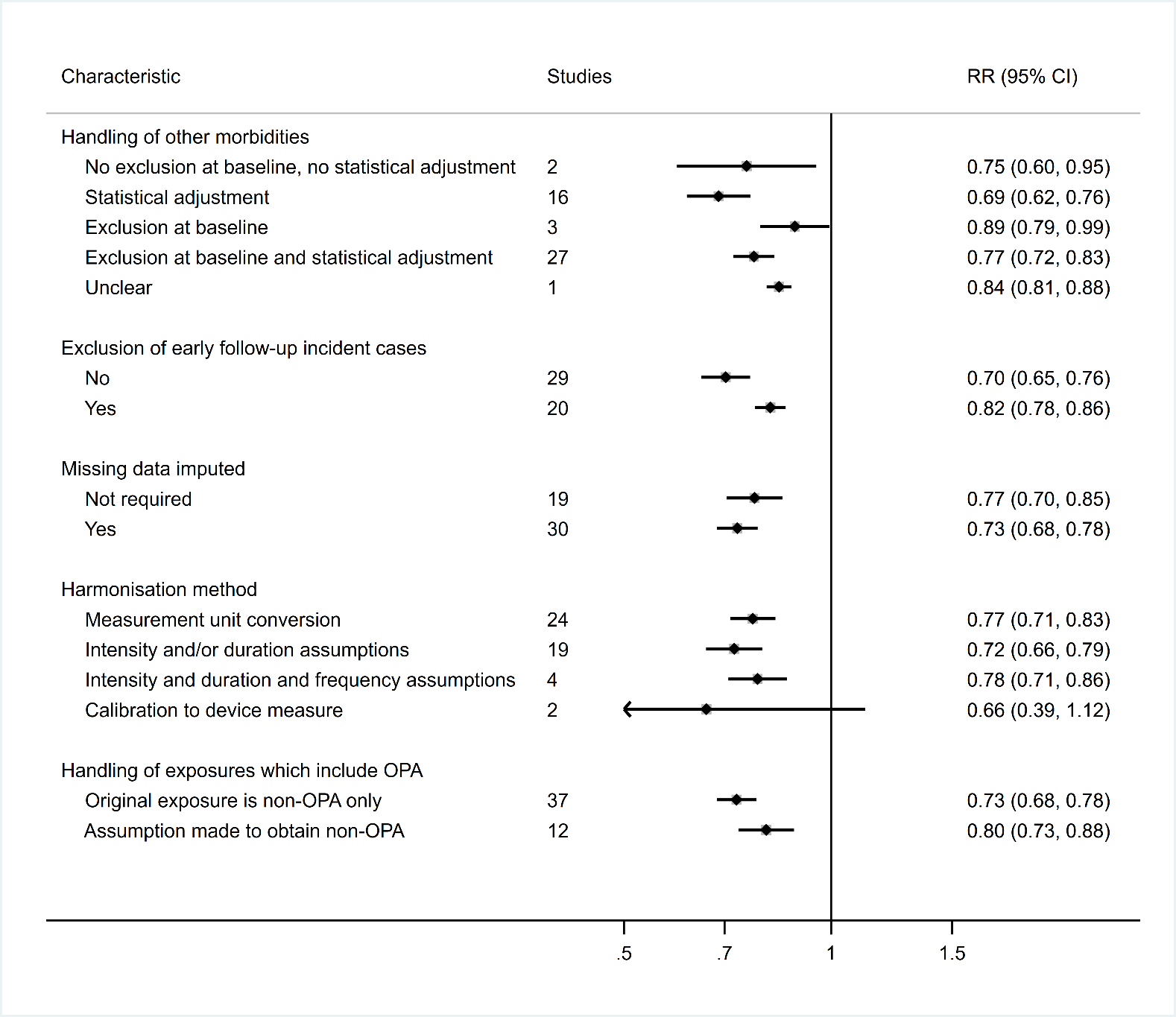

**Total CVD mortality**

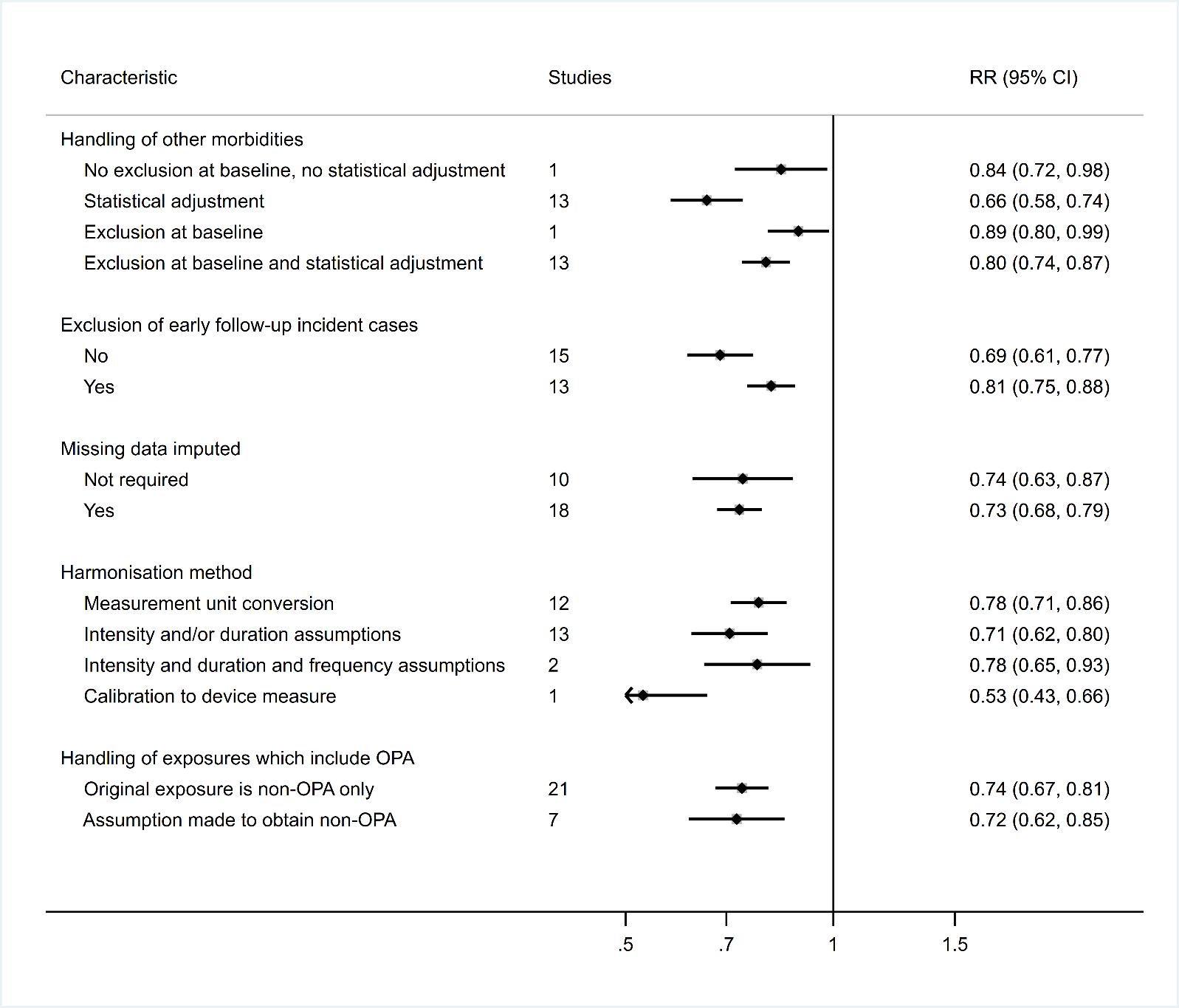

**Total cancer mortality**

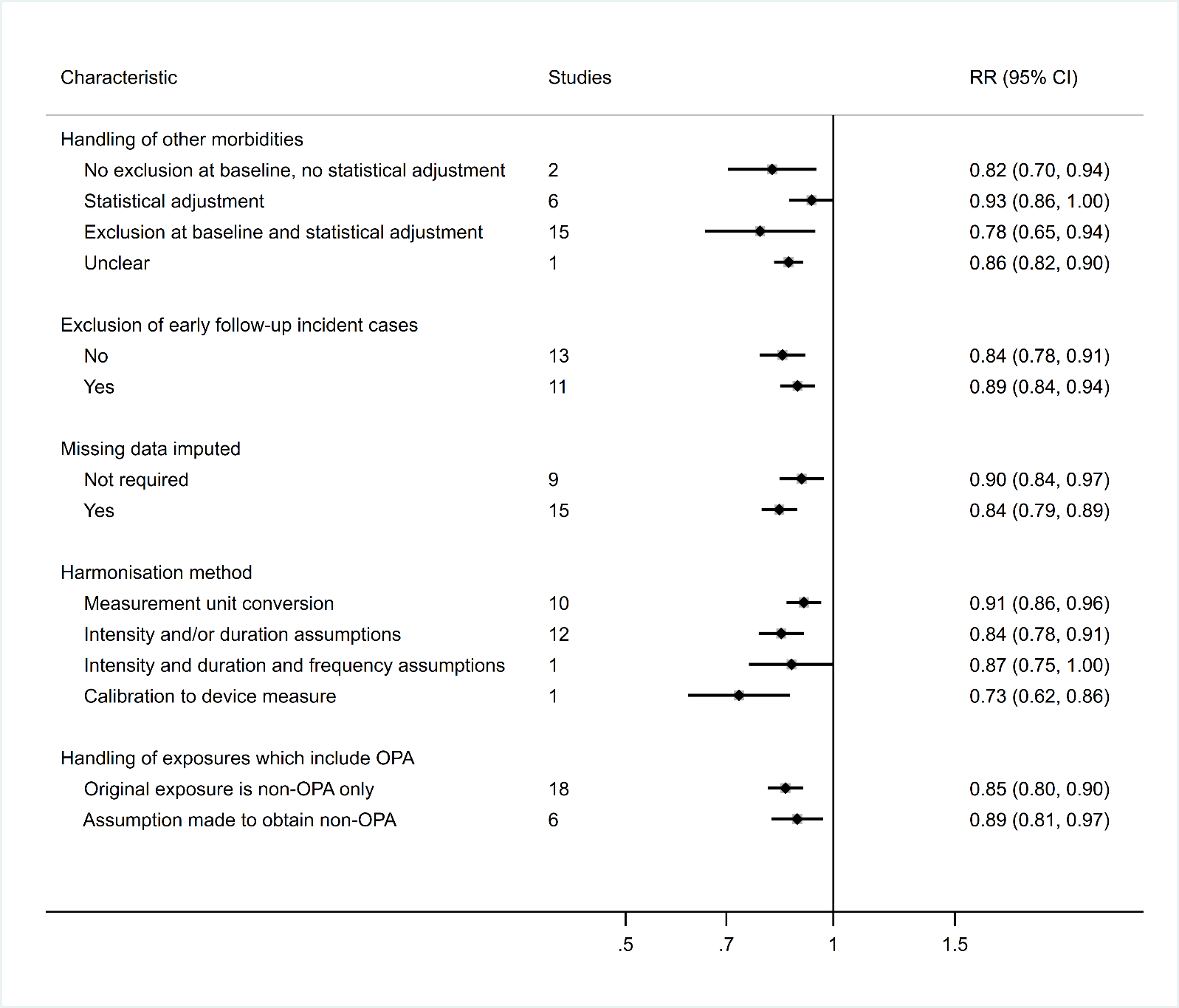

**Total CVD incidence**

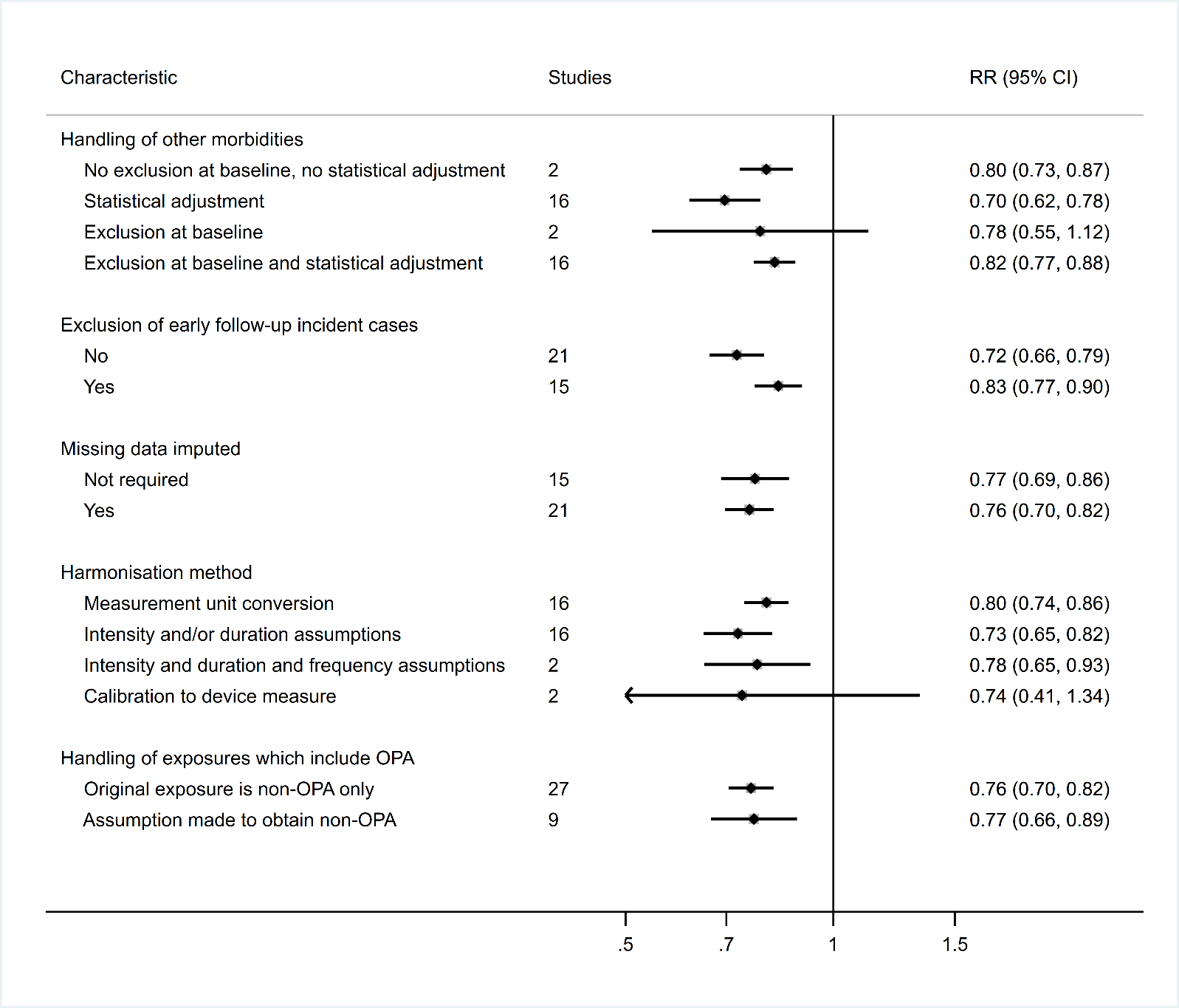

**Total cancer incidence**

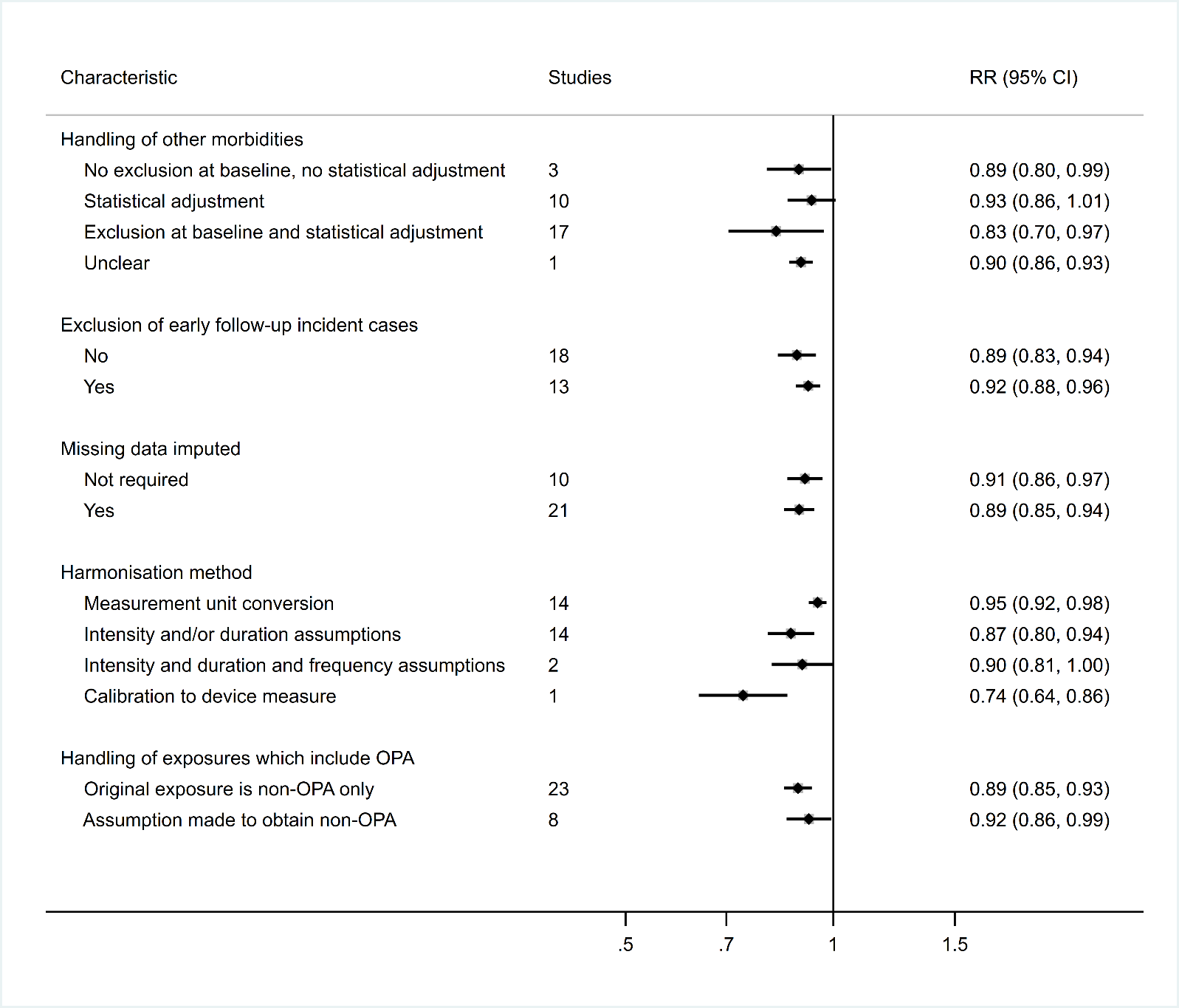

### **eResults 4: sensitivity analysis results**

| **Relative risk of all-cause mortality and incidence of cardiovascular diseases and cancers at three physical activity levels (in mMET-h/wk) after alternative placement of knots (0^th^, 42.5^th^, and 85^th^ percentile), alternative assumptions for physical activity intensity (1 mMET less to MPA and VPA) and session duration (0.5 hour rather than 0.75), and including only non-adiposity-adjusted models.** | | | | | | | | | | | |
| --- | --- | --- | --- | --- | --- | --- | --- | --- | --- | --- | --- |
| **Outcomes** | **Alternative knots position** | | |  | **Alternative PA intensity and duration assumptions** | | |  | **Non-adiposity-adjusted models only** | | |
|  | **4.375** | **8.75** | **17.5** |  | **4.375** | **8.75** | **17.5** |  | **4.375** | **8.75** | **17.5** |
|  | **RR (95% CI)** | **RR (95% CI)** | **RR (95% CI)** |  | **RR (95% CI)** | **RR (95% CI)** | **RR (95% CI)** |  | **RR (95% CI)** | **RR (95% CI)** | **RR (95% CI)** |
| Mortality |  |  |  |  |  |  |  |  |  |  |  |
| All-cause mortality | 0.78  (0.75 - 0.81) | 0.69  (0.65 - 0.74) | 0.66  (0.62 - 0.71) |  | 0.76  (0.72 - 0.79) | 0.67  (0.63 - 0.72) | 0.64  (0.60 - 0.69) |  | 0.69  (0.63 - 0.76) | 0.63  (0.57 - 0.71) | 0.60  (0.54 - 0.68) |
| Total CVD | 0.83  (0.79 - 0.87) | 0.73  (0.67 - 0.79) | 0.65  (0.60 - 0.71) |  | 0.77  (0.73 - 0.82) | 0.67  (0.61 - 0.74) | 0.63  (0.57 - 0.69) |  | 0.76  (0.68 - 0.84) | 0.64  (0.54 - 0.75) | 0.57  (0.47 - 0.69) |
| Total cancer | 0.91  (0.89 - 0.94) | 0.86  (0.82 - 0.89) | 0.82  (0.78 - 0.86) |  | 0.89  (0.86 - 0.92) | 0.84  (0.80 - 0.88) | 0.82  (0.79 - 0.86) |  | 0.86  (0.81 - 0.92) | 0.80  (0.74 - 0.87) | 0.77  (0.71 - 0.84) |
| CVD incidence |  |  |  |  |  |  |  |  |  |  |  |
| Total CVD | 0.85  (0.81 - 0.88) | 0.75  (0.70 - 0.80) | 0.68  (0.63 - 0.73) |  | 0.80  (0.76 - 0.84) | 0.70  (0.65 - 0.76) | 0.65  (0.60 - 0.71) |  | 0.78  (0.72 - 0.84) | 0.66  (0.59 - 0.75) | 0.60  (0.51 - 0.69) |
| Coronary heart disease | 0.88  (0.85 - 0.91) | 0.80  (0.76 - 0.85) | 0.74  (0.69 - 0.80) |  | 0.82  (0.78 - 0.87) | 0.77  (0.71 - 0.82) | 0.74  (0.68 - 0.80) |  | 0.84  (0.8 - 0.89) | 0.76  (0.71 - 0.82) | 0.71  (0.66 - 0.76) |
| Stroke | 0.88  (0.85 - 0.91) | 0.81  (0.76 - 0.85) | 0.76  (0.71 - 0.81) |  | 0.84  (0.81 - 0.88) | 0.80  (0.76 - 0.84) | 0.77  (0.72 - 0.84) |  | 0.86  (0.82 - 0.91) | 0.79  (0.74 - 0.85) | 0.75  (0.70 - 0.81) |
| Heart failure | 0.77  (0.63 - 0.94) | 0.66  (0.48 - 0.91) | 0.63  (0.42 - 0.94) |  | 0.90  (0.86 - 0.95) | 0.83  (0.76 - 0.91) | 0.75  (0.66 - 0.86) |  | 0.85  (0.79 - 0.93) | 0.77  (0.68 - 0.86) | 0.70  (0.63 - 0.78) |
| Cancer incidence |  |  |  |  |  |  |  |  |  |  |  |
| Total cancer | 0.94  (0.92 - 0.96) | 0.90  (0.87 - 0.93) | 0.85  (0.80 - 0.89) |  | 0.91  (0.89 - 0.94) | 0.87  (0.84 - 0.91) | 0.85  (0.81 - 0.89) |  | 0.89  (0.84 - 0.93) | 0.83  (0.78 - 0.90) | 0.81  (0.74 - 0.88) |
| Head and neck | 0.77  (0.63 - 0.94) | 0.66  (0.48 - 0.91) | 0.63  (0.42 - 0.94) |  | 0.72  (0.50 - 1.03) | 0.67  (0.45 – 1.00) | 0.63  (0.40 - 0.99) |  | 0.72  (0.59 - 0.88) | 0.63  (0.47 - 0.84) | 0.61  (0.41 - 0.91) |
| Myeloid leukemia | 0.85  (0.78 - 0.93) | 0.77  (0.67 - 0.88) | 0.74  (0.62 - 0.89) |  | 0.77  (0.60 - 0.99) | 0.75  (0.58 - 0.97) | 0.73  (0.54 - 0.97) |  | - | - | - |
| Myeloma | 0.84  (0.72 - 0.98) | 0.77  (0.62 - 0.96) | 0.76  (0.62 - 0.94) |  | 0.83  (0.72 - 0.96) | 0.75  (0.60 - 0.93) | 0.76  (0.60 - 0.97) |  | - | - | - |
| Gastric cardia | 0.89  (0.82 - 0.96) | 0.80  (0.70 - 0.92) | 0.73  (0.60 - 0.88) |  | 0.86  (0.77 - 0.96) | 0.78  (0.66 - 0.92) | 0.73  (0.60 - 0.90) |  | 0.90  (0.78 - 1.05) | 0.84  (0.66 - 1.08) | 0.79  (0.57 - 1.10) |
| Lung | 0.90  (0.88 - 0.93) | 0.85  (0.80 - 0.89) | 0.82  (0.74 - 0.91) |  | 0.87  (0.85 - 0.90) | 0.83  (0.79 - 0.87) | 0.84  (0.74 - 0.95) |  | 0.83  (0.78 - 0.88) | 0.76  (0.69 - 0.84) | 0.78  (0.66 - 0.91) |
| Liver | 0.90  (0.83 - 0.97) | 0.82  (0.72 - 0.94) | 0.75  (0.63 - 0.89) |  | 0.88  (0.79 - 0.97) | 0.82  (0.70 - 0.95) | 0.78  (0.65 - 0.93) |  | 0.89  (0.80 - 0.99) | 0.82  (0.68 - 0.98) | 0.76  (0.60 - 0.96) |
| Endometrial | 0.95  (0.92 - 0.99) | 0.91  (0.86 - 0.97) | 0.87  (0.79 - 0.95) |  | 0.93  (0.89 - 0.98) | 0.89  (0.82 - 0.97) | 0.86  (0.77 - 0.95) |  | 0.89  (0.84 - 0.95) | 0.81  (0.73 - 0.91) | 0.73  (0.62 - 0.86) |
| Colon | 0.97  (0.94 - 0.99) | 0.94  (0.89 - 0.99) | 0.91  (0.84 - 0.98) |  | 0.95  (0.91 - 0.99) | 0.92  (0.86 - 0.98) | 0.90  (0.83 - 0.97) |  | 0.97  (0.93 - 1.01) | 0.94  (0.87 - 1.02) | 0.92  (0.84 - 1.01) |
| Breast | 0.97  (0.96 - 0.99) | 0.95  (0.93 - 0.97) | 0.92  (0.89 - 0.95) |  | 0.96  (0.94 - 0.98) | 0.93  (0.90 - 0.97) | 0.91  (0.87 - 0.95) |  | 0.97  (0.94 – 1.00) | 0.95  (0.90 – 1.00) | 0.92  (0.86 - 0.98) |
| Bladder | 0.94  (0.86 - 1.02) | 0.91  (0.81 - 1.02) | 0.91  (0.81 - 1.02) |  | 0.92  (0.82 - 1.02) | 0.90  (0.81 - 1.01) | 0.89  (0.76 - 1.06) |  | - | - | - |
| *(continues)* | | | | | | | | | | | |

| **Relative risk of all-cause mortality and incidence of cardiovascular diseases and cancers at three physical activity levels (in mMET-h/wk) after assigning alternative knots position (0^th^, 42.5^th^, and 85^th^ percentile), using alternative assumptions for physical activity intensity (1 mMET less to MPA and VPA) and session duration (0.5 hour rather than 0.75), and including only non-adiposity-adjusted models (continuation).** | | | | | | | | | | | |
| --- | --- | --- | --- | --- | --- | --- | --- | --- | --- | --- | --- |
| **Outcomes** | **Alternative knots position** | | |  | **Alternative PA intensity and duration assumptions** | | |  | **Non-adiposity-adjusted models only** | | |
|  | **4.375** | **8.75** | **17.5** |  | **4.375** | **8.75** | **17.5** |  | **4.375** | **8.75** | **17.5** |
|  | **RR (95% CI)** | **RR (95% CI)** | **RR (95% CI)** |  | **RR (95% CI)** | **RR (95% CI)** | **RR (95% CI)** |  | **RR (95% CI)** | **RR (95% CI)** | **RR (95% CI)** |
| Rectum | 0.97  (0.93 - 1.01) | 0.95  (0.89 - 1.03) | 0.96  (0.87 - 1.05) |  | 0.95  (0.9 – 1.0) | 0.94  (0.87 - 1.01) | 0.96  (0.89 - 1.04) |  | 0.90  (0.84 - 0.97) | 0.87  (0.78 - 0.96) | 0.88  (0.79 - 0.98) |
| Esophageal | 0.98  (0.91 - 1.05) | 0.96  (0.85 - 1.08) | 0.95  (0.80 - 1.12) |  | 0.95  (0.85 - 1.07) | 0.93  (0.78 - 1.11) | 0.94  (0.77 - 1.13) |  | 0.91  (0.78 - 1.06) | 0.86  (0.67 - 1.11) | 0.85  (0.63 - 1.16) |
| Prostate | 1.00  (0.99 - 1.02) | 1.01  (0.98 - 1.03) | 1.01  (0.97 - 1.05) |  | 1.01  (0.99 - 1.03) | 1.01  (0.98 - 1.05) | 1.01  (0.96 - 1.05) |  | 1.01  (0.98 - 1.04) | 1.02  (0.97 - 1.07) | 1.02  (0.95 - 1.10) |
| Kidney | 1.02  (0.94 - 1.11) | 1.04  (0.91 - 1.19) | 1.05  (0.88 - 1.26) |  | 1.03  (0.90 - 1.16) | 1.04  (0.88 - 1.22) | 1.06  (0.84 - 1.33) |  | 0.88  (0.79 - 0.98) | 0.82  (0.71 - 0.96) | 0.79  (0.68 - 0.93) |
| mMET = marginal metabolic equivalent of task; RR = relative risk; 95% CI = 95% confidence interval. CVD = cardiovascular disease. | | | | | | | | | | | |

| **Potential impact fraction of physical activity on all-cause mortality and incidence of cardiovascular diseases and cancers at three physical activity levels (in mMET-h/wk) after alternative placement of knots (0^th^, 42.5^th^, and 85^th^ percentile), alternative assumptions for physical activity intensity (1 mMET less to MPA and VPA) and session duration (0.5 hour rather than 0.75), and including only non-adiposity-adjusted models.** | | | | | | | | | | | |
| --- | --- | --- | --- | --- | --- | --- | --- | --- | --- | --- | --- |
| **Outcomes** | **Alternative knots position** | | |  | **Alternative PA intensity and duration assumptions** | | |  | **Non-adiposity-adjusted models only** | | |
|  | **4.375** | **8.75** | **17.5** |  | **4.375** | **8.75** | **17.5** |  | **4.375** | **8.75** | **17.5** |
|  | **PIF (95% CI)** | **PIF (95% CI)** | **PIF (95% CI)** |  | **PIF (95% CI)** | **PIF (95% CI)** | **PIF (95% CI)** |  | **PIF (95% CI)** | **PIF (95% CI)** | **PIF (95% CI)** |
| Mortality |  |  |  |  |  |  |  |  |  |  |  |
| All-cause mortality | 10.11  (8.32 - 11.91) | 15.66  (13.16 - 18.15) | 18.75  (16.09 - 21.39) |  | 10.73  (8.88 - 12.62) | 17.08  (14.36 - 19.77) | 20.05  (17.01 - 23.04) |  | 13.98  (10.35 - 17.65) | 18.49  (14.15 - 22.8) | 21.99  (17.4 - 26.5) |
| Total CVD | 6.02  (4.42 - 7.69) | 12.25  (9.37 - 15.17) | 17.05  (13.92 - 20.19) |  | 8.00  (5.89 - 10.20) | 15.08  (11.52 - 18.68) | 18.66  (14.79 - 22.53) |  | 7.39  (4.41 - 10.67) | 13.83  (8.65 - 19.19) | 18.33  (12.26 - 24.43) |
| Total cancer | 3.68  (2.54 - 4.86) | 7.05  (5.10 – 9.00) | 9.29  (7.36 - 11.22) |  | 4.52  (3.13 - 5.94) | 7.71  (5.68 - 9.75) | 9.00  (7.20 - 10.80) |  | 7.22  (4.07 - 10.46) | 11.37  (7.09 - 15.68) | 13.73  (9.38 - 18.06) |
| CVD incidence |  |  |  |  |  |  |  |  |  |  |  |
| Total CVD | 5.29  (3.93 - 6.70) | 10.86  (8.43 - 13.31) | 15.42  (13.01 - 17.85) |  | 6.84  (5.10 - 8.63) | 13.16  (10.24 - 16.1) | 16.94  (14.07 - 19.82) |  | 6.43  (4.19 - 8.88) | 12.4  (8.41 - 16.54) | 17.18  (12.36 - 22.06) |
| Coronary heart disease | 5.51  (3.95 - 7.11) | 10.28  (7.61 - 12.96) | 13.58  (10.51 - 16.64) |  | 7.93  (5.58 - 10.32) | 12.13  (8.93 - 15.33) | 14.31  (10.43 - 18.14) |  | 6.26  (4.30 - 8.27) | 11.28  (8.23 - 14.33) | 14.97  (12.06 - 17.87) |
| Stroke | 5.53  (3.96 - 7.15) | 9.77  (7.31 - 12.24) | 12.37  (9.63 - 15.1) |  | 7.20  (5.33 - 9.10) | 10.40  (8.12 - 12.69) | 12.66  (8.46 - 16.77) |  | 5.13  (3.29 - 7.03) | 9.31  (6.51 - 12.14) | 12.25  (9.45 - 15.08) |
| Heart failure | 2.23  (0.80 - 3.89) | 4.63  (1.90 - 7.66) | 7.86  (4.31 - 11.68) |  | 2.36  (1.15 - 3.74) | 5.00  (2.49 - 7.75) | 9.43  (4.94 - 14.10) |  | 3.34  (1.34 - 5.78) | 6.25  (3.09 - 9.90) | 9.21  (5.86 - 12.84) |
| Cancer incidence |  |  |  |  |  |  |  |  |  |  |  |
| Total cancer | 2.53  (1.66 - 3.42) | 5.22  (3.55 - 6.90) | 7.80  (5.65 - 9.93) |  | 3.48  (2.33 - 4.66) | 6.56  (4.57 - 8.55) | 8.57  (6.31 - 10.81) |  | 5.67  (3.24 - 8.18) | 9.12  (5.51 - 12.76) | 11.12  (6.91 - 15.33) |
| Head and neck | 10.59  (2.10 - 19.96) | 17.46  (3.68 - 30.98) | 19.83  (1.23 - 36.55) |  | 13.82  (-1.07 - 28.5) | 18.21  (1.05 - 34.26) | 23.04  (2.16 - 41.14) |  | 11.66  (4.19 - 19.84) | 18.82  (6.67 - 30.81) | 20.07  (0.44 - 37.22) |
| Myeloid leukemia | 4.93  (0.68 - 9.70) | 7.08  (1.65 - 12.94) | 7.57  (2.23 - 13.33) |  | 6.67  (0.40 - 13.47) | 8.33  (1.91 - 15.25) | 10.22  (1.38 - 19.18) |  | - | - | - |
| Myeloma | 3.34  (0.90 - 6.26) | 6.76  (1.90 - 12.09) | 8.30  (1.91 - 14.99) |  | 3.71  (0.79 - 7.23) | 7.79  (1.75 - 14.37) | 6.57  (-0.43 - 14.07) |  | - | - | - |
| *(continues)* | | | | | | | | | | | |

| **Potential impact fraction of physical activity on all-cause mortality and incidence of cardiovascular diseases and cancers at three physical activity levels (in mMET-h/wk) after assigning alternative knots position (0^th^, 42.5^th^, and 85^th^ percentile), using alternative assumptions for physical activity intensity (1 mMET less to MPA and VPA) and session duration (0.5 hour rather than 0.75), and including only non-adiposity-adjusted models (continuation).** | | | | | | | | | | | |
| --- | --- | --- | --- | --- | --- | --- | --- | --- | --- | --- | --- |
| **Outcomes** | **Alternative knots position** | | |  | **Alternative PA intensity and duration assumptions** | | |  | **Non-adiposity-adjusted models only** | | |
|  | **4.375** | **8.75** | **17.5** |  | **4.375** | **8.75** | **17.5** |  | **4.375** | **8.75** | **17.5** |
|  | **PIF (95% CI)** | **PIF (95% CI)** | **PIF (95% CI)** |  | **PIF (95% CI)** | **PIF (95% CI)** | **PIF (95% CI)** |  | **PIF (95% CI)** | **PIF (95% CI)** | **PIF (95% CI)** |
| Gastric cardia | 1.95  (0.58 - 3.68) | 4.77  (1.58 - 8.57) | 6.77  (2.55 - 11.63) |  | 2.45  (0.64 - 4.78) | 5.40  (1.56 – 10.00) | 7.29  (2.34 - 13.03) |  | 1.13  (-0.42 - 3.29) | 3.17  (-1.19 - 8.63) | 5.12  (-1.81 - 13.09) |
| Lung | 4.39  (3.16 - 5.63) | 7.26  (5.10 - 9.4) | 8.54  (3.04 - 13.98) |  | 5.88  (4.05 - 7.37) | 9.56  (6.13 - 12.49) | 8.44  (-0.66 - 17.5) |  | 8.01  (4.69 - 11.23) | 12.27  (6.66 - 17.64) | 10.52  (0.77 - 20.21) |
| Liver | 3.49  (0.58 - 6.71) | 6.75  (1.44 - 12.31) | 10.27  (3.26 - 17.31) |  | 4.59  (0.90 - 8.59) | 8.37  (2.11 - 14.76) | 11.07  (3.45 - 18.63) |  | 4.62  (0.24 - 9.54) | 8.72  (0.79 - 16.97) | 12.86  (2.32 - 23.21) |
| Endometrial | 1.10  (0.26 - 2.02) | 2.62  (0.76 - 4.60) | 4.54  (1.88 - 7.28) |  | 1.50  (0.37 - 2.73) | 3.66  (1.07 - 6.38) | 5.91  (2.17 - 9.72) |  | 2.23  (0.89 - 3.77) | 5.66  (2.38 - 9.23) | 10.82  (4.99 - 16.76) |
| Colon | 0.70  (0.11 - 1.33) | 1.69  (0.33 - 3.12) | 2.96  (0.92 - 5.05) |  | 0.94  (0.19 - 1.74) | 2.12  (0.49 - 3.82) | 3.73  (1.24 - 6.25) |  | 0.45  (-0.15 - 1.11) | 1.19  (-0.32 - 2.79) | 2.29  (-0.06 - 4.70) |
| Breast | 0.69  (0.32 - 1.07) | 1.62  (0.79 - 2.46) | 3.23  (1.85 - 4.63) |  | 1.13  (0.53 - 1.75) | 2.44  (1.21 - 3.69) | 4.09  (2.35 - 5.83) |  | 0.79  (-0.05 - 1.67) | 1.78  (0.10 - 3.50) | 3.68  (1.45 - 5.94) |
| Bladder | 1.91  (-0.54 - 4.52) | 2.98  (-0.24 - 6.33) | 3.11  (0.02 - 6.35) |  | 2.72  (-0.64 - 6.23) | 3.70  (0.16 - 7.39) | 4.59  (-3.62 - 12.48) |  | - | - | - |
| Rectum | 0.77  (-0.15 - 1.75) | 1.43  (-0.45 - 3.36) | 0.52  (-1.74 - 2.82) |  | 1.30  (-0.09 - 2.74) | 2.07  (-0.49 - 4.65) | 0.02  (-2.60 - 2.67) |  | 2.07  (0.53 - 3.73) | 3.74  (0.90 - 6.66) | 2.69  (-0.19 - 5.66) |
| Esophageal | 0.68  (-0.95 - 2.68) | 1.38  (-2.06 - 5.32) | 1.57  (-3.05 - 6.67) |  | 1.02  (-1.34 - 3.88) | 1.87  (-2.65 - 6.95) | 1.58  (-3.44 - 7.13) |  | 1.86  (-1.08 - 5.71) | 3.7  (-2.48 - 10.93) | 3.96  (-4.07 - 12.86) |
| Prostate | -0.10  (-0.48 - 0.29) | -0.21  (-1.10 - 0.71) | -0.10  (-1.61 - 1.42) |  | -0.16  (-0.64 - 0.35) | -0.29  (-1.36 - 0.81) | -0.11  (-1.75 - 1.54) |  | -0.23  (-0.90 - 0.48) | -0.5  (-2.02 - 1.08) | -0.65  (-3.10 - 1.85) |
| Kidney | -0.65  (-3.69 - 2.81) | -1.23  (-6.39 - 4.34) | -1.91  (-9.20 - 5.63) |  | -0.95  (-5.25 - 3.85) | -1.78  (-8.56 - 5.40) | -3.06  (-15.89 - 9.34) |  | 5.61  (0.92 - 10.50) | 9.24  (2.27 - 16.16) | 11.73  (4.40 - 18.93) |
| mMET = marginal metabolic equivalent of task; PIF = potential impact fraction; 95% CI = 95% confidence interval. CVD = cardiovascular disease. | | | | | | | | | | | |
