## Supplementary material for "Non-occupational physical activity and risk of 22 cardiovascular disease, cancer, and mortality outcomes: a dose-response meta-analysis of large prospective studies": MOOSE checklist

**MOOSE Checklist for Meta-analyses of Observational Studies**

| **Item No** | **Recommendation** | | **Reported on Page No** |
| --- | --- | --- | --- |
| Reporting of background should include | | | |
| 1 | Problem definition | | 4 |
| 2 | Hypothesis statement | | - |
| 3 | Description of study outcome(s) | | 5 and eMethods 1 |
| 4 | Type of exposure or intervention used | | 5 and eMethods 1 |
| 5 | Type of study designs used | | 5 and eMethods 1 |
| 6 | Study population | | 5 and eMethods 1 |
| Reporting of search strategy should include | | | |
| 7 | Qualifications of searchers (eg, librarians and investigators) | | 5 |
| 8 | Search strategy, including time period included in the synthesis and key words | | 5 and eMethods 2 |
| 9 | Effort to include all available studies, including contact with authors | | eMethods 4 |
| 10 | Databases and registries searched | | 5 |
| 11 | Search software used, name and version, including special features used (eg, explosion) | | - |
| 12 | Use of hand searching (eg, reference lists of obtained articles) | | 5 |
| 13 | List of citations located and those excluded, including justification | | 9, eFigure 1 and OSF repository |
| 14 | Method of addressing articles published in languages other than English | | 5 |
| 15 | Method of handling abstracts and unpublished studies | | 5 |
| 16 | Description of any contact with authors | | 23 and eMethods 4 |
| Reporting of methods should include | | | |
| 17 | Description of relevance or appropriateness of studies assembled for assessing the hypothesis to be tested | | 5-7 |
| 18 | Rationale for the selection and coding of data (eg, sound clinical principles or convenience) | | 5-7 |
| 19 | Documentation of how data were classified and coded (eg, multiple raters, blinding and interrater reliability) | | 6 |
| 20 | Assessment of confounding (eg, comparability of cases and controls in studies where appropriate) | | 8 |
| 21 | Assessment of study quality, including blinding of quality assessors, stratification or regression on possible predictors of study results | | 8 |
| 22 | Assessment of heterogeneity | | 8 |
| 23 | Description of statistical methods (eg, complete description of fixed or random effects models, justification of whether the chosen models account for predictors of study results, dose-response models, or cumulative meta-analysis) in sufficient detail to be replicated | | 7-8 |
| 24 | Provision of appropriate tables and graphics | | Table 1, Figures 1-4, eResults 1-4, OSF repository and web interface |
| **Item No** | | **Recommendation** | **Reported on Page No** |
| Reporting of results should include | | | |
| 25 | Graphic summarizing individual study estimates and overall estimate | | Figures 1-4, Table 1 and web interface |
| 26 | Table giving descriptive information for each study included | | OSF repository |
| 27 | Results of sensitivity testing (eg, subgroup analysis) | | 18-19 and eResults 1-4 |
| 28 | Indication of statistical uncertainty of findings | | 18-21, Table 1, Figures 1-3 and eResults 1-4 |
| Reporting of discussion should include | | | |
| 29 | Quantitative assessment of bias (eg, publication bias) | | 18 |
| 30 | Justification for exclusion (eg, exclusion of non-English language citations) | | 5 |
| 31 | Assessment of quality of included studies | | 19 |
| Reporting of conclusions should include | | | |
| 32 | Consideration of alternative explanations for observed results | | 20-21 |
| 33 | Generalization of the conclusions (ie, appropriate for the data presented and within the domain of the literature review) | | 20-21 |
| 34 | Guidelines for future research | | 21 |
| 35 | Disclosure of funding source | | 22-23 |

*From*: Stroup DF, Berlin JA, Morton SC, et al, for the Meta-analysis Of Observational Studies in Epidemiology (MOOSE) Group. Meta-analysis of Observational Studies in Epidemiology. A Proposal for Reporting. *JAMA*. 2000;283(15):2008-2012. doi: 10.1001/jama.283.15.2008.
